# “What’s in a name?”: Using mpox as a case study to understand the importance of communication, advocacy, and information accuracy in disease nomenclature

**DOI:** 10.1101/2024.06.24.24309420

**Authors:** Erin N. Hulland, Marie-Laure Charpignon, Ghinwa Y. El Hayek, Angel N. Desai, Maimuna S. Majumder

## Abstract

Historically, many diseases have been named after the species or location of discovery, the discovering scientists, or the most impacted population. However, species-specific disease names often misrepresent the true reservoir; location-based disease names are frequently targeted with xenophobia; some of the discovering scientists have darker histories; and impacted populations have been stigmatized for this association. Acknowledging these concerns, the World Health Organization now proposes naming diseases after their causative pathogen or symptomatology. Recently, this guidance has been retrospectively applied to a disease at the center of an outbreak rife with stigmatization and misinformation: mpox (f.k.a. ‘monkeypox’). This disease, historically endemic to west and central Africa, has prompted racist remarks as it spread globally in 2022 in an epidemic ongoing today. Moreover, its elevated prevalence among men who have sex with men has yielded increased stigma against the LGBTQ+ community. To address these prejudicial associations, ‘monkeypox’ was renamed ‘mpox’ in November 2022.

We used publicly available data from Google Search Trends to determine which countries were quicker to adopt this name change—and understand factors that limit or facilitate its use. Specifically, we built regression models to quantify the relationship between ’mpox’ search intensity in a given country and the country’s type of political regime, robustness of sociopolitical and health systems, level of pandemic preparedness, extent of gender and educational inequalities, and temporal evolution of mpox cases through December 2023. Our results suggest that, when compared to ‘monkeypox’ search intensity, ’mpox’ search intensity was significantly higher in countries with any history of mpox outbreaks or higher levels of LGBTQ+ acceptance; meanwhile, ‘mpox’ search intensity was significantly lower in countries governed by leaders who had recently propagated infectious disease misinformation.

Among infectious diseases with stigmatizing names, mpox is among the first to be revised retrospectively. While the adoption of a given disease name will be context-specific—depending in part on its origins and the affected subpopulations—our study provides generalizable insights, applicable to future changes in disease nomenclature.

## Introduction

Historically, diseases have often been named after the animal that first presented with the pathogen (e.g., monkeypox, swine flu, bird flu); the location of first diagnosis or report (e.g., Spanish flu, Marburg virus, Ebola Zaire virus); or the discovering scientist or affected patient’s name (e.g., Hodgkin’s lymphoma, Alzheimer’s disease, Chagas disease). Since the release of specific guidelines in 2015, the World Health Organization (WHO) recommends avoiding such historical practices and instead favors focusing on symptomatology or the causative pathogen when naming new diseases.^1^ This recommendation is responsive to rising xenophobia and avoidance related to location-centric names (e.g., anti-Mexican sentiment following the “Mexican swine flu” moniker for the 2009 H1N1 pandemic^2^), culling of animal populations wrongly believed to be reservoirs for a disease (e.g., pig culling in Egypt due to the 2009 H1N1 “swine flu” pandemic^3^), and occasionally, the prejudicial past of a discovering scientist (e.g., Rett syndrome, named after an Austrian neurologist who was a member of the Nazi party in his youth^4^). In some cases, a disease name may not even accurately reflect its origins. This latter scenario is best illustrated by the Spanish flu, which likely originated in North America, but was first reported in Spain due to limited communication during World War I.^5^

The magnitude of the COVID-19 pandemic shined a bright light on pervasive xenophobia and racism tied to the origins of its causative pathogen—SARS-CoV-2—and on the importance of appropriate risk communication. While authors of scientific publications generally followed the new WHO guidance in naming the disease caused by the novel coronavirus (SARS-CoV-2), prominent figures in politics and the media have not similarly followed suit, often emphasizing the origins of COVID-19 by using inflammatory language to refer to the virus. One such example is former President Trump’s reference to SARS-CoV-2 as the “Chinese virus” in 2020.^6^ Given the use of such stigmatizing language in tweets and press briefings, both online anti-Asian sentiment and physical violence against Asians and Asian Americans rose dramatically.^6–12^

While the COVID-19 pandemic raged on, a simultaneous mpox (f.k.a. ‘monkeypox’) outbreak spread to over 50 countries in 2022.^13^ Previously, mpox was endemic to west and central Africa, with only a few sporadic outbreaks recorded in other countries up until 2022—during which an unprecedented epidemic, comprised of multiple country-level outbreaks, unfolded.^14–16^ While previous mpox outbreaks in endemic regions did not exhibit sexual transmission, this mode of transmission was observed for the first time during the 2022 epidemic.^17^ Given the prevalence of sexual transmission during this epidemic, similarities with the early HIV/AIDS epidemic were drawn, leading to increased stigmatization of gay men in particular.^18–20^ Because of the epidemic’s unprecedented global reach, the presentation and naming of the disease were more visible than ever before. The name ‘monkeypox’ presented not only a misnomer, as monkeys are highly unlikely to be the reservoir for mpox, but also perpetuated an offensive stereotype of African populations, among whom the bulk of mpox infections had previously occurred.^21–23^ Following a call by the WHO to rename the disease in August 2022, the name ‘monkeypox’ was formally changed to ‘mpox’ on November 28, 2022, allowing for one year of simultaneous use of the two names.^24,25^

The transition from ‘monkeypox’ to ‘mpox’ is the first known example of a formal name change for an infectious disease in the Internet Age, designed to explicitly address prejudice and misinformation. It echoes former experiences with AIDS in the early 1980s, when the disease was referred to as “Gay-Related Immune Deficiency” or GRID.^26,27^ Similar to mpox, which was named after its first identification in a captive cynomolgus monkey,^28^ this ‘GRID’ was named after its first presentation in gay men in Los Angeles in 1980,^29^ resulting in tremendous stigmatization of the LGBTQ+ community. Ultimately, in August 1982, the Centers for Disease Control introduced a non-stigmatizing name for this novel disease: Acquired Immune Deficiency Syndrome (AIDS). Nevertheless, the damage associated with the initial name had long-term repercussions—and this stigma persists today.^30^ As such, the 2015 WHO recommendation endeavors to avoid the use of stigmatizing language for newly-named diseases, yet no formal and sufficiently broad guidance has been provided to date for retroactively changing existing disease names en masse, despite countless examples of existing disease names with prejudicial connotations that would require such intervention.^1^ In the absence of formal retrospective naming guidelines, this event not only provides an interesting case study but also creates a precedent for the WHO and its national counterparts to build upon.

Despite this formal guidance, the global adoption of the new term has been inconsistent. As such, our goal was to understand which factors influenced whether a country was a timely adopter of the name change from ‘monkeypox’ to ‘mpox’. In this study, we leveraged Google Search Trends (GST) to examine country-level search intensity of the terms ‘mpox’ and ‘monkeypox’ between November 2022 and December 2023, the time period over which the WHO recommended the dual use of both ‘mpox’ and ‘monkeypox’. Previously, GST has been used to study infectious diseases such as seasonal influenza, SARS-CoV-2, and measles, among others—from predicting and monitoring their spread, to understanding spatiotemporal variation in health-seeking behaviors, discovering new disease symptoms via search trends, developing tailored risk communication plans, and more.^31–40^

Our main analytic goal was to understand the drivers of adoption of the term ‘mpox’ at the country-level. To that end, we developed two key outcomes: (1) a binary variable reflecting whether search intensity was higher for ‘mpox’ or ‘monkeypox’ during our study period; and (2) a continuous variable defined as the fraction of the search intensity for ‘mpox’ and ‘monkeypox’ attributable to ‘mpox’. We constructed regression models to associate these outcomes with static country-level factors such as type of political regime, robustness of sociopolitical and health systems, level of pandemic preparedness, extent of gender and educational inequalities, temporal evolution of confirmed local mpox cases during the 2022 epidemic, and distribution of mpox cases since discovery. Through this study, we hope to provide insights not only into country-level factors that were most conducive to the public’s adoption of the destigmatized name—as reflected by the intensity of Google search queries—but also into improving adoptions during future disease name changes.

## Methods

### Data sourcing and pre-processing

#### Google Search Trends

We obtained the trajectory of Google search intensity for ‘mpox’ and ‘monkeypox’ over time through Google Search Trends (GST), a publicly-available online platform that provides anonymized, categorized, and aggregated data. We included 184 countries with available GST data on either term throughout the study period, out of 249 distinct countries and territories captured by the platform overall. Given that English is currently the *lingua franca* in science, thus serving as the language of disease communication and nomenclature, we relied on Google Search queries made in English in our primary analysis. However, we acknowledge the inherent power imbalance of Anglocentrism.^41–46^ Therefore, in an effort to also understand the dynamics of public interest in ‘mpox’ versus ‘monkeypox’ in countries where English is not the primary language, we considered the five other official WHO / United Nations (UN) languages: Arabic, Chinese, French, Russian, and Spanish. We report the results of this sensitivity analysis, aimed at further understanding the intensity of searches that users made in their native language, in the Appendix.

GST returns the proportion of Internet search activity accounted for by each search term in a given location (e.g., metropolitan area, state, country) at a given point in time (e.g., day, week). This quantity, ranging between 0% and 100%, is defined as the number of term-specific searches in that location at that particular point in time divided by the total number of searches over that same region and timespan.^47,48^ By collecting data for both search terms simultaneously (i.e., ‘mpox’ and ‘monkeypox’), we were able to compare their respective search intensity on the same spatiotemporal scale for the period ranging from November 1, 2022 to December 1, 2023. In doing so, our intent was to capture one full year of Google search intensity data following the WHO announcement on November 28, 2022 and thus study the progressive adoption of the term ‘mpox’ by the general public.

#### Outcome variables

We derived two outcome variables defined at the country level. Both variables were based on the average search intensity for the terms ‘mpox’ and ‘monkeypox’ in a given country over the study period. The first outcome, a **binary** variable, captured whether a country had greater average search intensity for ‘mpox’ than for ‘monkeypox’ (1 if yes, 0 otherwise), allowing us to distinguish ‘mpox’ adopters from non-adopters among the 184 countries considered in our study. The second outcome, a **continuous** variable, was defined as the fraction of search intensity attributed to ‘mpox’ over the study period (i.e., the ratio of the search intensity for ‘mpox’ over the total search intensity for ‘mpox’ and ‘monkeypox’ combined, expressed as a percentage value); we refer to this variable hereafter as the *‘mpox’ search proportion.* While we defined two distinct outcomes to test the sensitivity of our findings to the use of metrics with different granularity (i.e., binary vs continuous), our overall analytic goal was to identify the consistent drivers of ‘mpox’ adoption; thus, our results are presented and interpreted jointly.

#### Covariates

In this study, we sought to identify country-level factors contributing to adoption of the term ‘mpox’ around the globe. To achieve this goal, we compiled a suite of 18 publicly-available country-level covariates to model their associations with ‘mpox’ search intensity (Table 1). Given high LGBTQ+ stigma during the 2022 mpox epidemic, we included a country-level measure of LGBTQ+ acceptance, called the Global Acceptance Index (GAI).^49^ The GAI is a continuous variable ranging from 0 to 10. Since pandemics have become increasingly politicized, as exemplified by COVID-19 in the United States and Brazil,^50,51^ we accounted for various measures of a country’s current political context. Specifically, we relied on variables previously used to understand the COVID-19 pandemic,^52,53^ including the type of political regime of the country (four categories: closed autocracy, electoral autocracy, electoral democracy, or liberal democracy);^54^ the Electoral Democracy Index, a continuous variable ranging from 0 to 1 that acts as a proxy for free and fair elections as well as voting rights;^54^ the Liberal Democracy Index, a continuous variable ranging from 0 to 1 that explores election freedom, civil liberties, and limits to executive power;^54^ and the Corruption Perceptions Index, a continuous variable ranging from 0 to 100 that measures the perceived levels of corruption in the public sector.^55^

Because the health system and its practitioners are often the first point of contact for disease presentation and reporting, we were also interested in understanding whether healthcare access and quality of care in a given country influenced the public’s perception and knowledge of a disease, and ultimately, its nomenclature. To this end, we included the Healthcare Access and Quality Index, a continuous variable ranging from 0 to 100, as a covariate.^56^ Moreover, we capitalized on the 2021 Global Health Security Index (GHSI)—a composite index comprising 171 variables across 37 indicators in total—to measure a country’s preparedness to detect, prevent, and respond to future infectious disease outbreaks. Given the rise in emerging and re-emerging infections, we deemed it relevant to characterize each country’s approach to health communication, their strategies to mitigate disease spread, and their targeted efforts to protect at-risk populations. To this end, we focused on three GHSI components related to risk communications.^57^ In particular, we considered the GHSI aggregate risk communications score, as well as two related—but more specific—measures: the inclusiveness of a country’s risk communication plans and the prior use of infectious disease misinformation by a country’s government. The first was captured by GHSI question 3.5.1b, which focuses on an explicit intention to communicate with hard-to-reach populations based on language, rurality, and media access. The second was captured by GHSI question 3.5.2b, which characterizes whether a country’s senior leaders (i.e., presidents or prime ministers) had shared misinformation or disinformation about infectious diseases in the previous two years. Additionally, we leveraged four GHSI components related to inequalities in mobile phone and Internet access, both between genders within a given country and among countries. In the first case, the ratio of access to mobile phones and Internet between males and females in a given country provides an indicator of gender-based disparities; in the second case, country-level indicators of mobile phone and Internet access capture Internet and technology penetration in a country. In both cases, limited access to mobile phones and the Internet ultimately influence who is able to use Internet search platforms like Google Search.

We also accounted for disparities in country wealth, reasoning that the average income level is associated with the average level of English proficiency of the population and thus their likelihood of searching for English terms.^58^ Additionally, given that the 2022 mpox epidemic was heavily concentrated in high- and upper-middle-income countries, we wanted to control for the distribution of country wealth in our analyses.^59^ For these reasons, we included the gross domestic product (GDP) per capita as a covariate.^60^ Reasoning that a country’s average educational attainment would be associated with the public’s propensity to search for ‘mpox’, we similarly explored between-country educational disparities using the mean number of years of schooling among women as a proxy; we specifically chose education levels among women rather than among men to capture a greater heterogeneity among countries due to inequitable access to education for women around the world. Last, we considered the impact that the recent 2022 mpox epidemic may have had on the adoption of ‘mpox’ by the public. More specifically, to assess the relative timing of the epidemic in a given country, we calculated the number of days between the first confirmed mpox case worldwide (in the United Kingdom on May 6, 2022)^61^ and that country’s first confirmed case. In addition, we created a binary indicator of whether a country had ever had a confirmed case of mpox in humans or mammals since it was first discovered in humans in 1970, in order to gauge previous levels of exposure to the disease and its name.^59,62,63^

Given varying patterns of missingness in the covariates across the 184 countries included in the analysis (Appendix Figure 2), we used a multiple imputation approach with predictive mean matching over ten imputations. We considered three different combinations of data selection and imputation methods: (1) selecting countries with no missing data and performing a complete-case analysis with these resulting 154 countries; (2) selecting countries that either had complete data or were missing only the GAI and imputing this variable where necessary, yielding a sample size of 166 countries; and (3) imputing all variables with missing data, resulting in a total of 184 countries. Results presented in the main text capitalize on the fully imputed covariate dataset (i.e., Approach 3); results relying on the two other imputation strategies (i.e., Approaches 1 and 2) are presented in the Appendix. Transformations made to the covariate data in pursuit of normalizing the distribution are available in the Appendix Section 3.1.

#### Modeling

Following data extraction and pre-processing, we built univariable and multivariable regression models separately for each of our two outcomes. The binary outcome—whether a country was an adopter (1) or not (0)—was modeled using logistic generalized linear models with time-invariant covariates. The continuous outcome—‘mpox’ search proportion—was modeled using generalized linear regression models with the same time-invariant covariates. First, we evaluated the strength of the simple association between each outcome of interest and each of the 18 covariates using univariable models. We retained all variables significant at p<0.1, removed those that were highly collinear, and subsequently implemented multivariable models adjusted for the remaining subset (Table 1). We accounted for uncertainty over the ten iterations of the imputation process by drawing from a multivariable normal distribution; then, we summarized across all ten draws using the median and the 2.5th and 97.5th percentiles to obtain a 95% uncertainty interval.

#### Sensitivity analyses

In addition to our secondary analyses based on Google searches made in languages other than English (Appendix Figures 5 and 6), we conducted six sensitivity analyses to assess the robustness of our results to data and modeling decisions. First, we compared our main results with those emanating from the two alternative imputation strategies described above, namely the complete case analysis (Approach 1-Appendix Figures 7 and 8) and the imputation of only the GAI variable (Approach 2 - Appendix Figures 9 and 10); descriptive statistics for these imputation approaches are also visible in Table 1. Second, we evaluated the impact of restricting our dataset to only the countries that ever had a confirmed case of mpox in order to understand the role of exposure on the adoption of the term ‘mpox’ by the public (Appendix Figures 11 and 12). Third, we considered the impact of using a more stringent significance threshold, at p<0.05 rather than p<0.1, for covariate inclusion into our multivariable regression models (Appendix Figure 13). Fourth, we used a Bonferroni correction to account for post-hoc testing of multiple hypotheses at once (Appendix Figures 14 and 15). Finally, we implemented a Tobit model, well-suited for zero-inflated data, to address a large proportion of zeros in our continuous outcome measuring the ‘mpox’ search proportion (Appendix Figure 16).

### Data availability and role of the funder

All analyses were conducted using R version 4.3.1^64^. This research was supported in part by the National Institute of General Medical Sciences, National Institutes of Health (R35GM146974); the National Science Foundation (SES2200228, SES2230083, & IIS2229881); and the MIT-Harvard Broad Institute Eric & Wendy Schmidt Center. The funding sources had no involvement in the study design; in the collection, analysis, and interpretation of data; in the writing of the report; or in the decision to submit the paper for publication.

## Results

In analyzing the intensity of Google searches for ‘mpox’ and ‘monkeypox’ across over 180 countries worldwide, we identified three consistent country-level factors that were significantly associated with adoption of the term ‘mpox’: any history of mpox in the country, greater LGBTQ+ acceptance, and senior leaders who did not recently propagate infectious disease misinformation; our results were robust to the imputation strategy chosen. We present key summary statistics (e.g., median and interquartile range or IQR) for all measured covariates, both pre- and post-imputation, in Table 1. Additionally, we examine a cross-comparison of the three imputation strategies described in the Methods section to assess their relative impact on our model covariates and outcomes (Table 1).

**Table 1:**
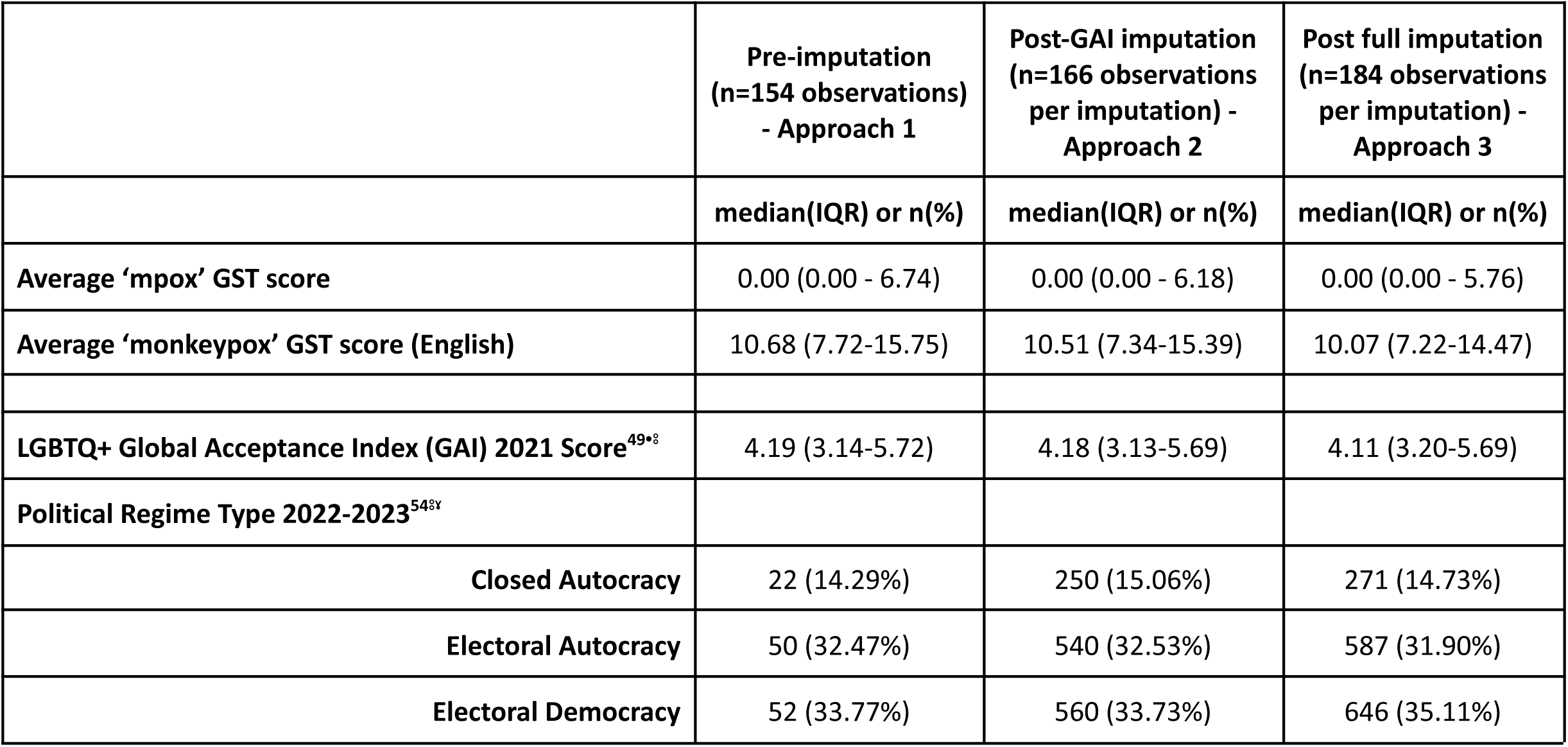
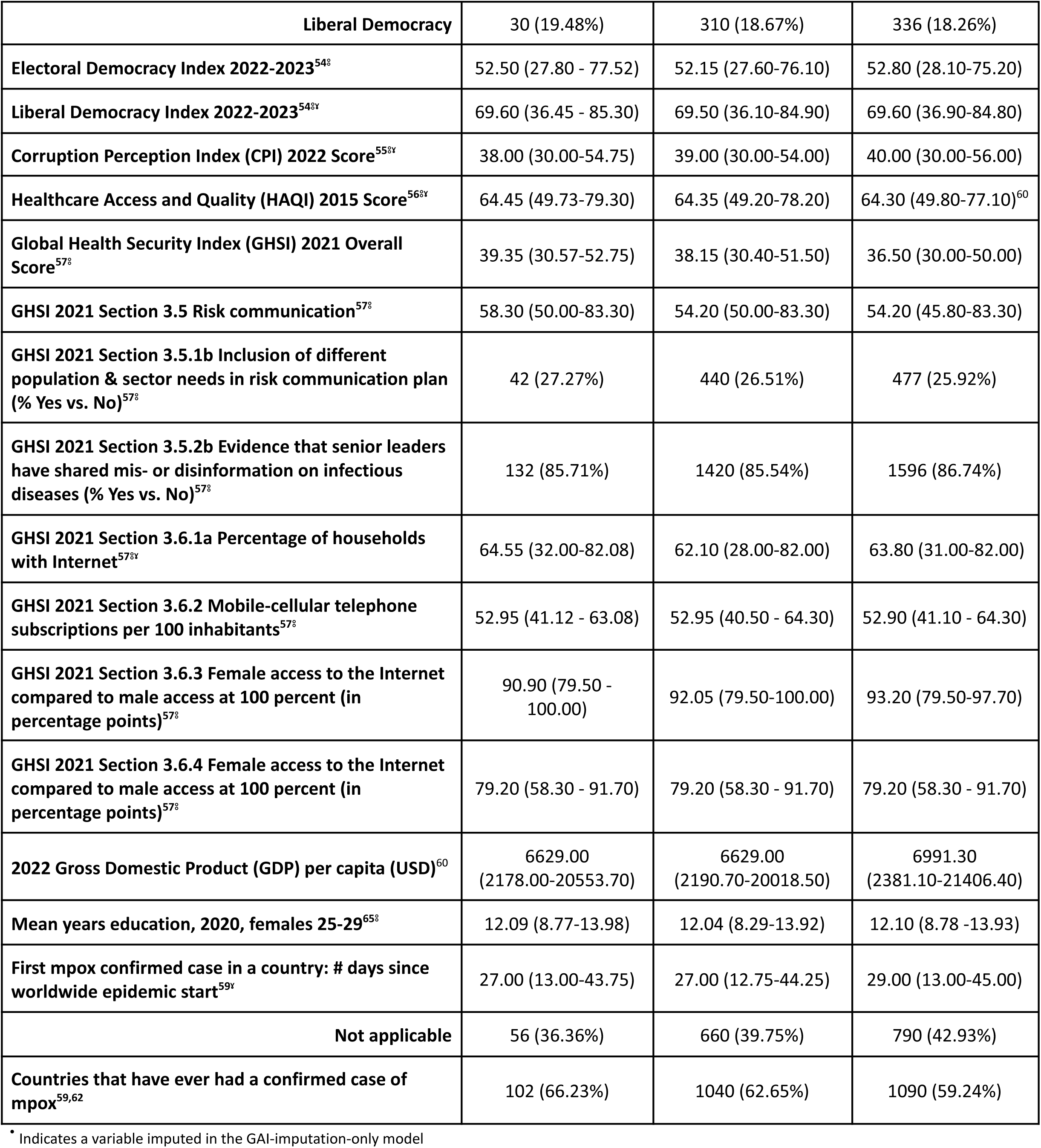
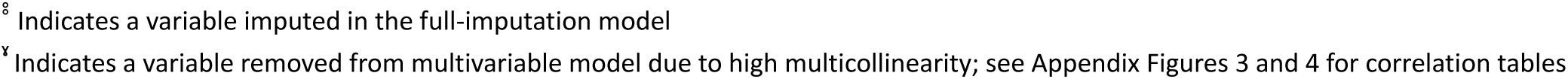
Key covariates pre-imputation, after imputing the LGBTQ+ Global Acceptance Index only, and after imputing all data.

|  | Pre-imputation<br>(n=154 observations)<br>- Approach 1 | Post-GAI imputation<br>(n=166 observations<br>per imputation) -<br>Approach 2 | Post full imputation<br>(n=184 observations<br>per imputation) -<br>Approach 3 |
| --- | --- | --- | --- |
|  | median(IQR) or n(%) | median(IQR) or n(%) | median(IQR) or n(%) |
| Average ‘mpox’ GST score | 0.00 (0.00 - 6.74) | 0.00 (0.00 - 6.18) | 0.00 (0.00 - 5.76) |
| Average ‘monkeypox’ GST score (English) | 10.68 (7.72-15.75) | 10.51 (7.34-15.39) | 10.07 (7.22-14.47) |
| LGBTQ+ Global Acceptance Index (GAI) 2021 Score<sup>49*</sup> | 4.19 (3.14-5.72) | 4.18 (3.13-5.69) | 4.11 (3.20-5.69) |
| Political Regime Type 2022-2023<sup>54**</sup> |  |  |  |
| Closed Autocracy | 22 (14.29%) | 250 (15.06%) | 271 (14.73%) |
| Electoral Autocracy | 50 (32.47%) | 540 (32.53%) | 587 (31.90%) |
| Electoral Democracy | 52 (33.77%) | 560 (33.73%) | 646 (35.11%) |
| **Liberal Democracy** | 30 (19.48%) | 310 (18.67%) | 336 (18.26%) |
| **Electoral Democracy Index 2022-2023<sup>54§</sup>** | 52.50 (27.80 - 77.52) | 52.15 (27.60-76.10) | 52.80 (28.10-75.20) |
| **Liberal Democracy Index 2022-2023<sup>54§y</sup>** | 69.60 (36.45 - 85.30) | 69.50 (36.10-84.90) | 69.60 (36.90-84.80) |
| **Corruption Perception Index (CPI) 2022 Score<sup>55§y</sup>** | 38.00 (30.00-54.75) | 39.00 (30.00-54.00) | 40.00 (30.00-56.00) |
| **Healthcare Access and Quality (HAQI) 2015 Score<sup>56§y</sup>** | 64.45 (49.73-79.30) | 64.35 (49.20-78.20) | 64.30 (49.80-77.10)<sup>60</sup> |
| **Global Health Security Index (GHSI) 2021 Overall Score<sup>57§</sup>** | 39.35 (30.57-52.75) | 38.15 (30.40-51.50) | 36.50 (30.00-50.00) |
| **GHSI 2021 Section 3.5 Risk communication<sup>57§</sup>** | 58.30 (50.00-83.30) | 54.20 (50.00-83.30) | 54.20 (45.80-83.30) |
| **GHSI 2021 Section 3.5.1b Inclusion of different population & sector needs in risk communication plan (% Yes vs. No)<sup>57§</sup>** | 42 (27.27%) | 440 (26.51%) | 477 (25.92%) |
| **GHSI 2021 Section 3.5.2b Evidence that senior leaders have shared mis- or disinformation on infectious diseases (% Yes vs. No)<sup>57§</sup>** | 132 (85.71%) | 1420 (85.54%) | 1596 (86.74%) |
| **GHSI 2021 Section 3.6.1a Percentage of households with Internet<sup>57§y</sup>** | 64.55 (32.00-82.08) | 62.10 (28.00-82.00) | 63.80 (31.00-82.00) |
| **GHSI 2021 Section 3.6.2 Mobile-cellular telephone subscriptions per 100 inhabitants<sup>57§</sup>** | 52.95 (41.12 - 63.08) | 52.95 (40.50 - 64.30) | 52.90 (41.10 - 64.30) |
| **GHSI 2021 Section 3.6.3 Female access to the Internet compared to male access at 100 percent (in percentage points)<sup>57§</sup>** | 90.90 (79.50 - 100.00) | 92.05 (79.50-100.00) | 93.20 (79.50-97.70) |
| **GHSI 2021 Section 3.6.4 Female access to the Internet compared to male access at 100 percent (in percentage points)<sup>57§</sup>** | 79.20 (58.30 - 91.70) | 79.20 (58.30 - 91.70) | 79.20 (58.30 - 91.70) |
| **2022 Gross Domestic Product (GDP) per capita (USD)<sup>60</sup>** | 6629.00 (2178.00-20553.70) | 6629.00 (2190.70-20018.50) | 6991.30 (2381.10-21406.40) |
| **Mean years education, 2020, females 25-29<sup>65§</sup>** | 12.09 (8.77-13.98) | 12.04 (8.29-13.92) | 12.10 (8.78 -13.93) |
| **First mpox confirmed case in a country: # days since worldwide epidemic start<sup>59y</sup>** | 27.00 (13.00-43.75) | 27.00 (12.75-44.25) | 29.00 (13.00-45.00) |
| **Not applicable** | 56 (36.36%) | 660 (39.75%) | 790 (42.93%) |
| **Countries that have ever had a confirmed case of mpox<sup>59,62</sup>** | 102 (66.23%) | 1040 (62.65%) | 1090 (59.24%) |

\* Indicates a variable imputed in the GAI-imputation-only model
<sup>8</sup> Indicates a variable imputed in the full-imputation model
<sup>y</sup> Indicates a variable removed from multivariable model due to high multicollinearity; see Appendix Figures 3 and 4 for correlation tables

In Figure 1a, nine variables were significantly associated with our binary outcome in our univariable model; meanwhile, when considering our continuous outcome—’mpox’ search proportion—there were 16 significant variables in the univariable model (Figure 2a). When we included only those variables with univariate significance p<0.1 in the two multivariable models, we observed three consistent significant relationships across our two outcomes: (1) a positive relationship with countries who had ever reported an mpox case, either as part of the 2022 epidemic or in a prior outbreak; (2) a positive relationship with LGBTQ+ GAI score; and (3) a negative relationship with countries with senior leaders who had recently employed misinformation about any infectious disease (Figures 1b and 2b).

**Figure 1 -.**
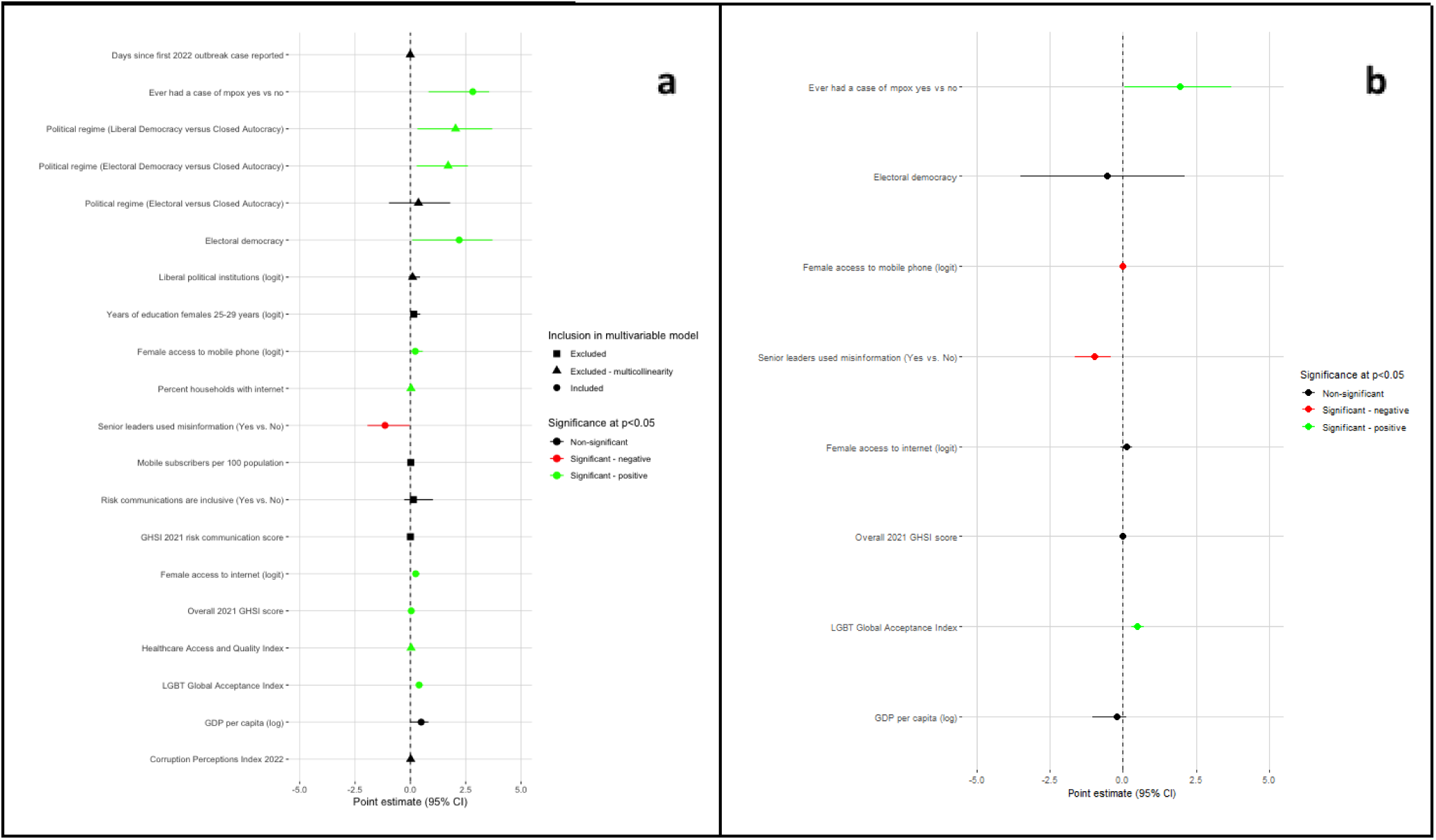
Country-level factors associated with ‘mpox’ adopter countries (binary outcome) in univariable (a) and multivariable (b) models. Variables that were statistically significantly associated with the binary outcome of interest at alpha = 0.1 in the corresponding univariable models were included in the multivariable model. Highly collinear variables were removed before fitting the multivariable model. Triangle markers in panel (a) denote covariates removed due to high multicollinearity; square markers indicate those covariates removed due to non-statistical significance at alpha = 0.1 in their corresponding univariable model; circle marker indicate covariates included in the multivariable model.

**Figure 2 -.**
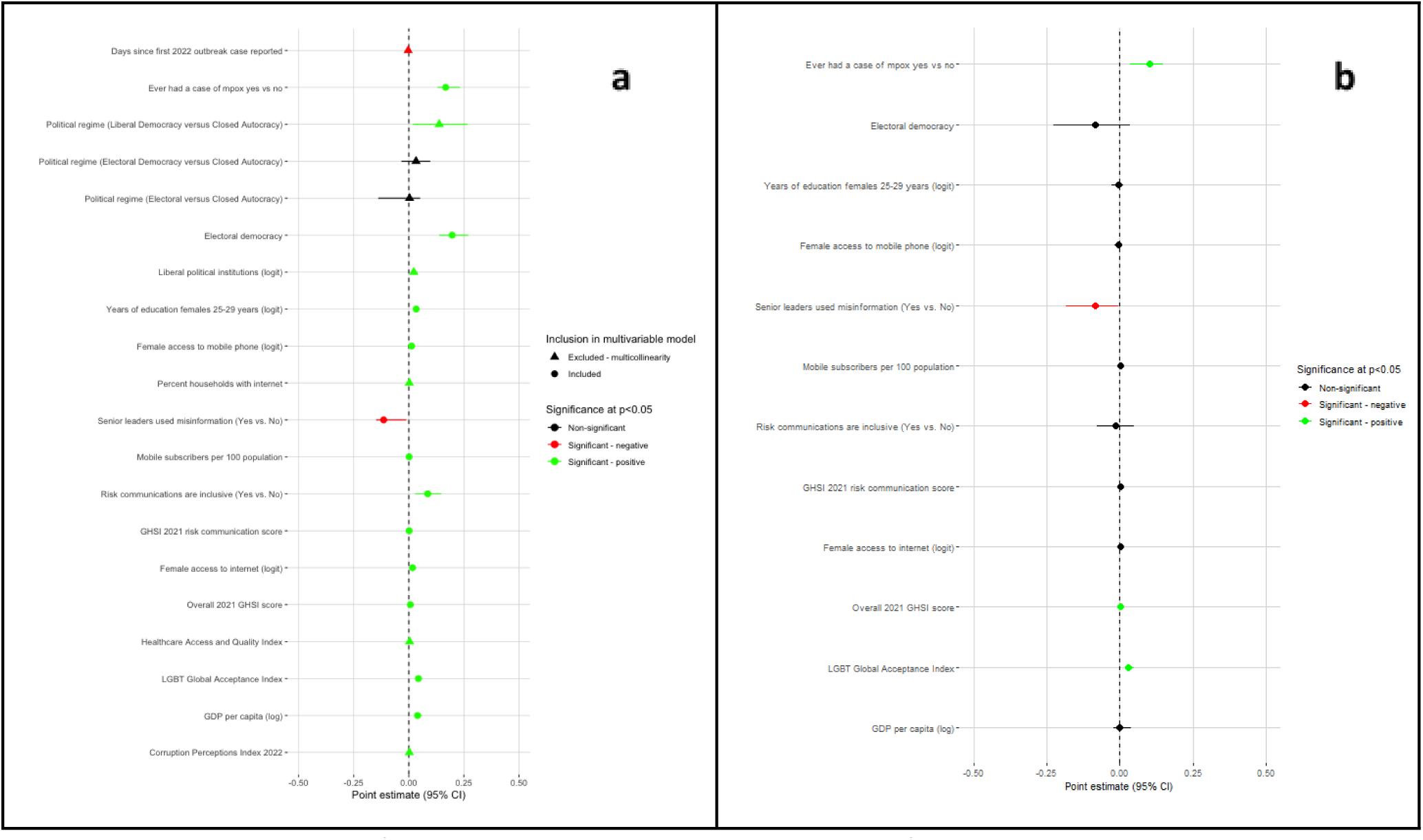
Country-level factors associated with ‘mpox’ search proportion (continuous outcome) in univariable (a) and multivariable (b) models. Variables that were statistically significantly associated with the continuous outcome of interest at alpha = 0.1 in the corresponding univariable models were included in the multivariable model. Highly collinear variables were removed before fitting the multivariable model. Triangle markers in panel (a) denote covariates removed due to high multicollinearity; square markers indicate those covariates removed due to non-statistical significance at alpha = 0.1 in their corresponding univariable model; circle marker indicate covariates included in the multivariable model.

## Discussion

Our study suggests that countries with higher LGBTQ+ acceptance, as measured by the GAI, were more likely to adopt the term ‘mpox’ over the term ‘monkeypox’ in the year following the WHO recommendation of the name change. This result suggests that countries where the public is more accepting of queer communities were also more likely to demonstrate a more thoughtful use of destigmatizing language when conducting Google searches. Importantly, this finding was consistent across all sensitivity analyses we conducted, emphasizing its robustness to the choice of imputation strategy and model type. In contrast to historic mpox outbreaks that predominantly affected west and central Africa and had no reported sexual transmission, the 2022 multi-country epidemic of mpox has largely been categorized as a disease among the gay, bisexual, and other men who have sex with men (GBMSM) community.^66,67^ As a result, the GBMSM community has faced backlash and stigma—in addition to the fear of mpox itself.^19,20,68,69^ Therefore, our study underscores the need to increase awareness and acceptance of marginalized populations. Indeed, as exemplified in previous outbreaks—most notably HIV/AIDS— stigma associated with sexual transmission not only impacts quality of life, but can also limit a person’s access to care and reduce their propensity to accurately report their behaviors. These negative repercussions can in turn lead to delayed treatment, poorer health outcomes, and sustained disease propagation.^70–73^ Thus, in order to increase awareness and acceptance of marginalized populations going forward, strong risk communications combating bigotry, prejudice, and stigma are critical early in infectious disease crises.^74^

Another strong correlate of ‘mpox’ search intensity was countries that had ever had a confirmed case of mpox—in humans and other mammals alike. We hypothesize that people living in places with previous cases had increased exposure to news and social media about mpox, increasing awareness of the new name. Previous research about the impact of news media on infectious diseases—including mpox, H1N1, Ebola, and COVID-19—has demonstrated that increased exposure to quality information through news and social media can increase vaccine uptake,^75,76^ reduce disease transmission,^77^ and encourage protective behaviors.^78–80^ Our work reflects the need for awareness campaigns during infectious disease crises, emphasizing proper nomenclature as well as providing high-quality information on disease spread and populations most at-risk. Future research evaluating media communications regarding the mpox name change may provide valuable insights into how the news media referred to the disease over time; whether there were any political, geographical, or temporal trends; and what messaging at what time point corresponded to higher uptake of ‘mpox’. In doing so, more specific guidance can be generated for future media campaigns on infectious disease name changes.

We identified a third strong correlate of ‘mpox’ adoption: in countries where senior leaders had recently propagated infectious disease misinformation, adoption was significantly lower. Such misinformation has been increasingly identified as a barrier to vaccines,^81–84^ seeking timely care,^81,85,86^ and positive behavior change; ^81,87^ has eroded trust in governments and authorities;^88–90^ and has ultimately worsened health outcomes.^86,89,91^ During the COVID-19 pandemic, misinformation was employed by politicians in the US and abroad to promote the use of untested medicine such as Ivermectin, Azithromycin, Chloroquine and Hydroxychloroquine;^92–94^ the avoidance of many public health guidelines like vaccination or mask wearing;^95–97^ and dangerous practices like ingesting or injecting bleach,^98,99^ resulting in surges in product purchases, price hikes, and in some cases, serious injury or death.^92,98–101^ More recently, a sentiment analysis of tweets suggested that mpox misinformation contributed to heightened stigma of LGBTQ+ populations, resulting in increased violence, harassment and isolation.^74^ Several strategies have been employed to combat the growing threat of misinformation: emphasizing quality sources of information; capitalizing on trusted leaders, officials, and scientists to convey accurate information; correcting and calling out misinformation; and increasing frequency of both proactive, accurate messaging and correcting inaccurate information.^102^ Currently, consequences incurred by leaders or politicians who spread misinformation vary by country and region. First, they may face financial and legal consequences, including penalties such as fines and, in some cases, imprisonment. Second, social media platforms themselves may take action and either restrict user privileges (e.g., retweeting, sharing, or posting) or ban such leaders outright; yet many countries still struggle to walk the line between action and censorship.^103,104^ While elected officials who propagate misinformation should undoubtedly be sanctioned, no one course of action is likely to suit every country. In the future, researchers should — in collaboration with dedicated counter-misinformation oversight committees, when possible — seek to understand how effective each strategy is at reducing the spread of misinformation in a given country, providing evidence to guide local decision making.

### Limitations

There are several limitations to this study. First, our main analysis relied on English as the *lingua franca* of scientific communication and our sensitivity analysis considered the five additional UN languages. These five languages—while widely used—account for only a small fraction of all the languages spoken across the world; as such, we are likely missing search terms for ‘monkeypox’ expressed in other languages, resulting in an incomplete capture of Google search interest in the disease globally. While we acknowledge the diversity of languages worldwide and its importance in public health, around 4 billion people speak at least one of the 6 UN languages as a primary or secondary language, representing about half of the world’s population.^105,106^

Second, consistent with the WHO nomenclature announcement, we only searched the single terms ‘mpox’ and ‘monkeypox’ rather than the composite terms ‘mpox virus’ and ‘monkeypox virus’, which remains the official taxonomic name. However, given the boolean methodology used by GST, data collected for the term ‘monkeypox’ or ‘mpox’ constitute a superset that would also account for the terms ‘monkeypox virus’ and ‘mpox virus’.^107^

Third, at the time of our analysis, foreign language data for two countries—Ecuador and Madagascar—were unavailable. To address this limitation, we attempted a counterfactual sensitivity analysis for the binary outcome, generating all permutations of the possible second-language findings and observed largely similar results to our main analyses—suggesting the robustness of our key findings (Appendix Section 3.7 and Figure 17).

Fourth, this study employs Internet searches conducted by users via Google Search alone, thus excluding those searches attributable to other search engines. Future research examining searches for ‘mpox’ and ‘monkeypox’ on other search engines (e.g., Bing, DuckDuckGo, Yahoo) would thus provide valuable comparative insights. Importantly, over 90% of Internet users across the globe use Google as their primary search engine—with the exception of China and Russia, suggesting GST are capturing the majority of global Internet searches.^108,109^

Last, we employed a suite of 18 covariates in our regression models, with varying degrees of missingness. We used multiple imputation with as many complete variables as possible, but acknowledge differences in resulting sample sizes and covariate data distributions between the complete-case analysis (Approach 1, n=154), the GAI-only imputation analysis (Approach 2, n=166), and the full imputation analysis (Approach 3, n=184). The most notable difference observed was GDP. Since this covariate was available for all 184 included countries, we could contrast its overall distribution to that in the analyses restricted to 154 (Approach 1) and 166 (Approach 2) countries, respectively.

Specifically, we observed a lower median GDP in both the GAI-only imputation analysis and the complete-case analysis. This result suggests that health system quality, pandemic preparedness, gender equitability, and social acceptance, which are all correlated with GDP, were likely not missing at random. Nonetheless, all of our sensitivity analyses of imputation strategies yielded similar main results, reinforcing the robustness of our conclusions.

## Conclusions

We have seen a surge of zoonotic disease transmission, outbreaks, and pandemics in this century so far; this trend is likely to continue with increasing population connectivity and mobility, deforestation and development encroaching on zoonotic reservoirs, and climate change influencing transmission. As such, the potential for stigmatizing language and xenophobia, as well as their downstream impacts and outcomes, are becoming increasingly visible. We need to plan now for ways to counter this stigma, and learning from those countries most successful in instituting ‘mpox’ in lieu of ‘monkeypox’ is a good way to start.

## Data Availability

All data in this analysis are openly available and provided via citations.

## Acknowledgements

We would like to thank Paula Rodriguez Diaz and Robyn Correll for their assistance in understanding ‘mpox’ trends in South America and the translation of ‘monkeypox’ in Spanish; Nilufar Qahorova for her assistance in translating ‘monkeypox’ to Russian; and Zhanzhan Zhao in her assistance in translating ‘monkeypox’ to Chinese.

## Section 1: Abbreviations

CPI: Corruption Perception Index
GAI: Global Acceptance Index
GBMSM: Gay, Bisexual, and other Men who have Sex with Men
GDP: Gross Domestic Product
GST: Google Search Trends
GHSI: Global Health Security Index
HAQI: Healthcare Access and Quality Index
LGBTQ+: Lesbian, Gay, Bisexual, Transgender, Queer or Questioning, intersex, asexual, and more
MSM: Men who have Sex with Men
WHO: World Health Organization
UI: Uncertainty Interval

## Section 2: Additional Tables

**Table 1:** UN official languages by country, excluding English (n=65)

| Country | Official language | Second official language |
| --- | --- | --- |
| Algeria | Arabic |  |
| Argentina | Spanish |  |
| Bahrain | Arabic |  |
| Belgium | French |  |
| Bolivia | Spanish |  |
| Burkina Faso | French |  |
| Burundi | French |  |
| Cameroon | French |  |
| Canada | French |  |
| Chile | Spanish |  |
| China | Chinese |  |
| Colombia | Spanish |  |
| Congo | French |  |
| Costa Rica | Spanish |  |
| Cuba | Spanish |  |
| Djibouti | Arabic | French |
| Dominican Republic | Spanish |  |
| Democratic Republic of the Congo | French |  |
| Egypt | Arabic |  |
| El Salvador | Spanish |  |
| Equatorial Guinea | French | Spanish |
| France | French |  |
| Gabon | French |  |
| Guatemala | Spanish |  |
| Guinea | French |  |
| Haiti | French |  |
| Honduras | Spanish |  |
| Hong Kong | Chinese |  |
| Iraq | Arabic |  |
| Côte d'Ivoire | French |  |
| Jordan | Arabic |  |
| Kazakhstan | Russian |  |
| Kuwait | Arabic |  |
| Kyrgyzstan | Russian |  |
| Lebanon | Arabic |  |
| Libya | Arabic |  |
| Mali | French |  |
| Mauritania | Arabic |  |
| Mexico | Spanish |  |
| Morocco | Arabic |  |
| Nicaragua | Spanish |  |
| Niger | French |  |
| Oman | Arabic |  |
| Palestine | Arabic |  |
| Panama | Spanish |  |
| Paraguay | Spanish |  |
| Peru | Spanish |  |
| Qatar | Arabic |  |
| Russia | Russian |  |
| Rwanda | French |  |
| Saudi Arabia | Arabic |  |
| Senegal | French |  |
| Seychelles | French |  |
| Singapore | Chinese |  |
| Spain | Spanish |  |
| Sudan | Arabic |  |
| Switzerland | French |  |
| Syria | Arabic |  |
| Togo | French |  |
| Tunisia | Arabic |  |
| United Arab Emirates | Arabic |  |
| Uruguay | Spanish |  |
| Vanuatu | French |  |
| Venezuela | Spanish |  |
| Yemen | Arabic |  |

This table shows the 65 countries where one of the five UN / WHO languages (other than English) are spoken. In our model, we substituted the English term for the UN language term for these 65 countries, but kept the English term for the remaining 119 countries, resulting in a total sample size of 184 countries for the analysis.

**Table 2:**
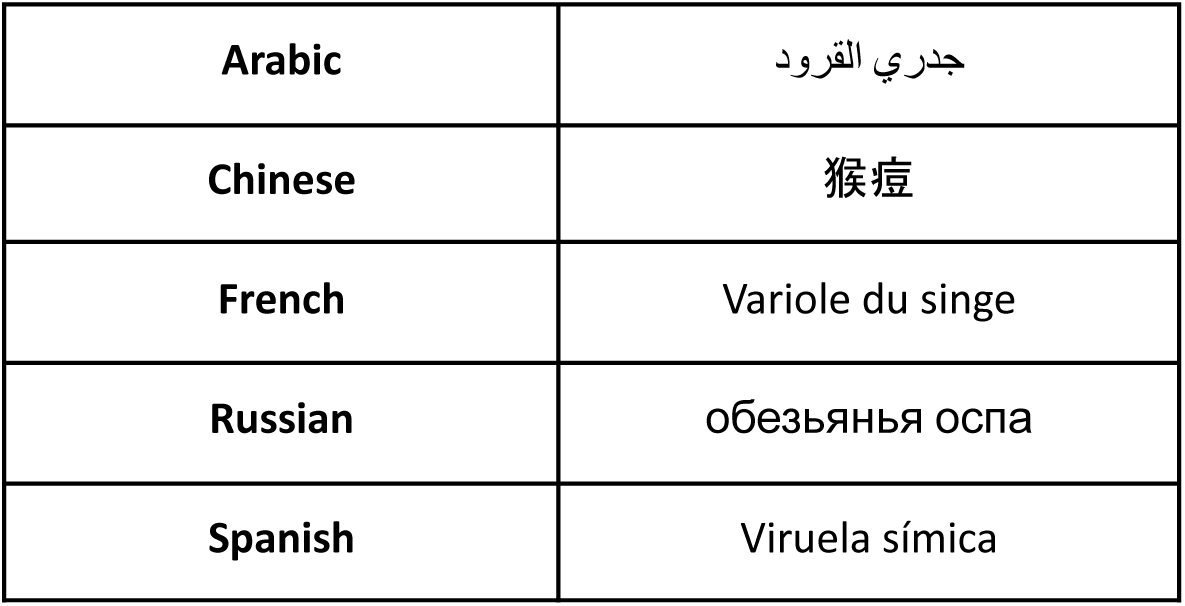
Monkeypox search term in 5 UN languages (excluding English)

## Section 3: Additional Methods

### Section 3.1: Transforming independent covariates

Several of our variables required cleaning and updating. For GDP per capita, we took the most recent year of data available for each country, between 2014 and 2022. 92.6% percent of countries had data from 2022, while 7.4% used data from older years. For data on historic mpox outbreaks, we used an occurrence database of all autochthonous mpox cases in humans or animals between 1970s and 2019, extracted from published literature;^1^ we additionally added the US, UK, and Israel which had imported mpox cases during these years. We then turned to recent outbreak data acquired from Our World in Data.^2^ We used these data to create two variables: 1) the number of days between the start of the 2022 outbreak and first case in a country with countries not included in the 2022 outbreak had day hard coded as 999 to avoid erroneously imputing the day since it is not truly missing, but then were removed for all univariable analyses; and 2) a binary indication of ever having had a case of mpox (either in the 2022 outbreak or earlier) or not ever. Due to high collinearity with ever having had a case of mpox, days since outbreak start was not included in any multivariable models.

We observed five highly skewed variables: four left-skewed, and one right skewed. Our right-skewed variable - GDP per capita - was transformed using a natural-log function. Our four left-skewed variables - Mean female education in years, Gender gap access internet, Gender gap access access mobile phone, the Liberal Democracy Index - were transformed using a logit function, after normalizing and scaling to a 0 to 1 scale. Because we were much more interested in the general trend of the data rather than specific predictions for each country, we wanted to maximize our available data. As such, where we used a logit transformation, we performed an empirical logit transformation for any variables with values of either 0 or 1: we added a minimal value ε defined as 0.5/n (where n=184 observations) to both the numerator and denominator during the logit transformation to the observations with a value of 0 or 1.^3^ Two of our variables, both from the 2021 GHSI framework - Inclusive risk communications and leaders sharing misinformation - had scores of either 0 or 100 with no other variation, so we dichotomized this variable into 0 = NO and 100 = YES.

### Section 3.2: Creating outcome variables

To generate our Google Search Trend data into our two outcome variables we performed a few cleaning and analytic functions. First, where data were listed as “<1“, we changed these to values of 0. For our binary variable, we first calculated the mean score for each mpox, monkeypox (or monkeypox in a second language) separately for each country and search term over the entire period. If the mean mpox search score was greater than the mean search score for monkeypox, we assigned a value of 1, otherwise we assigned a value of 0. For our continuous outcome, we calculated the percentage of searches that were mpox by day using the function *daily mpox search score/(daily mpox search score+daily monkeypox search score).* Where scores were NA (where both the numerator and denominator were 0), we imputed the value as 0. We then took the mean percent value by country over the entire time period to generate our continuous outcome variable per country

### Section 3.3: Additional methods for listwise deletion

For our imputation analyses, we performed ten imputations and summarized our data across these ten imputations. In order to match our imputed analysis, we took 10 multivariate normal draws for the listwise deletion analysis (Approach 1) as well, and summarized using the median and 95% uncertainty interval.

### Section 3.4: Foreign language search model

In order to better understand native-language searches versus English search, we considered all countries that had a WHO / UN official language as one of the official languages in the country (Table 1).^4^ We then performed the same search procedure, including both ‘mpox’ and the foreign-language term for ‘monkeypox’ (Table 2) simultaneously over the same study duration: 11/01/2022 through 12/01/2023. It is important to note that ‘mpox’ was chosen to be more language-agnostic and thus there was no translation done for this term. In two countries with multiple official languages (Djibouti and Equatorial Guinea), we considered both languages but ultimately used the first official language given no search data for the term ‘mpox’ in either country, resulting in no differences in either our binary or our continuous outcome using any of the selected languages. Results can be seen in Figures 5 and 6 below.

### Section 3.5: Tobit model

We identified a large number of zero scores for the percentage of all searches (mpox and monkeypox) that were mpox, due to a large number of places that did not have any mpox searches during our analysis period. While we also consider a binary outcome variable, which is one way to handle many zeros, we wanted to understand the role of model selection on our continuous outcome results. As such, we constructed a Tobit model using the VGLM function in R and a Tobit family with a lower bound of 0. For our univariable analysis, we removed the regime categorical variable due to lack of convergence; this was always removed from the multivariable analysis due to high collinearity. We present the results of the Tobit model on the fully imputed dataset and using a multivariable threshold of 0.1 in Figure 16.

### Section 3.6: Bonferonni correction

To account for multiple hypothesis testing, we performed a post-hoc Bonferroni correction. Since we were investigating 18 variables, we used a correction factor of α/18 for our uncertainty intervals. We re-ran all analyses, using the fully imputed data and an inclusion threshold at 0.1/18=0.00556. Results for the binary and continuous outcomes are visible in Figures 14 and 15 below, respectively.

### Section 3.7: Counterfactual analysis for Ecuador and Madagascar

For our sensitivity analysis looking at the five UN languages (section 3.4), data were not available at the time of analysis for Ecuador and Madagascar, despite being on the list of countries with official second languages (Table 1). As such, to analyze the impact of these missing data, we created counterfactual scenarios for our binary outcome to test how our results would vary. In the English-language analysis, Ecuador had a value of 1 for mpox searches being greater than monkeypox searches, while Madagascar had a value of 0, indicating mpox searches were fewer than monkeypox searches; in fact, Madagascar had no searches for mpox at all. We tested out counterfactuals where both Ecuador and Madagascar were 0 (mpox searches < monkeypox searches), Ecuador and Madagascar were both 1 (mpox searches > monkeypox searches), and the complete opposite of the English analysis where Madagascar was 1 and Ecuador was 0. We re-ran all of our regression models (univariable and multivariable) using the fully imputed data set with inclusion at 0.1. Results can be seen in Figure 17 below.

## Section 4: Additional Figures

**Figure 1:**
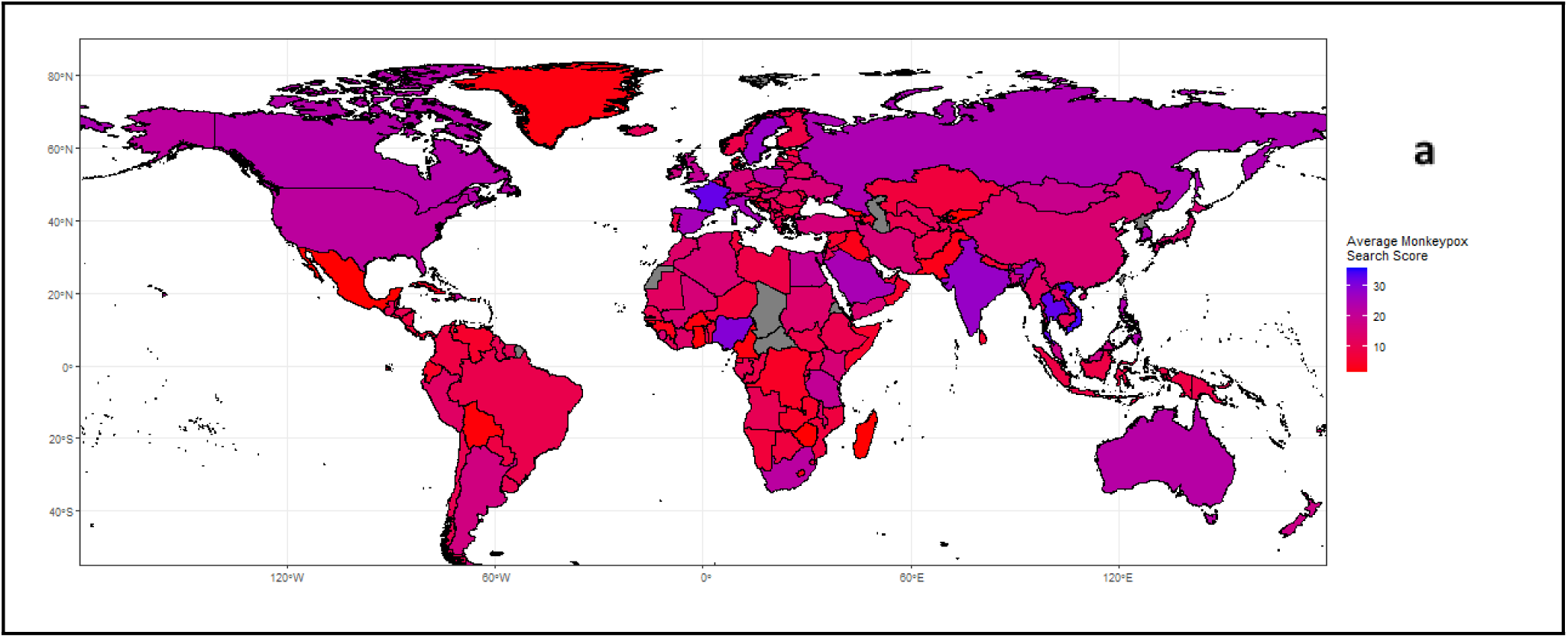
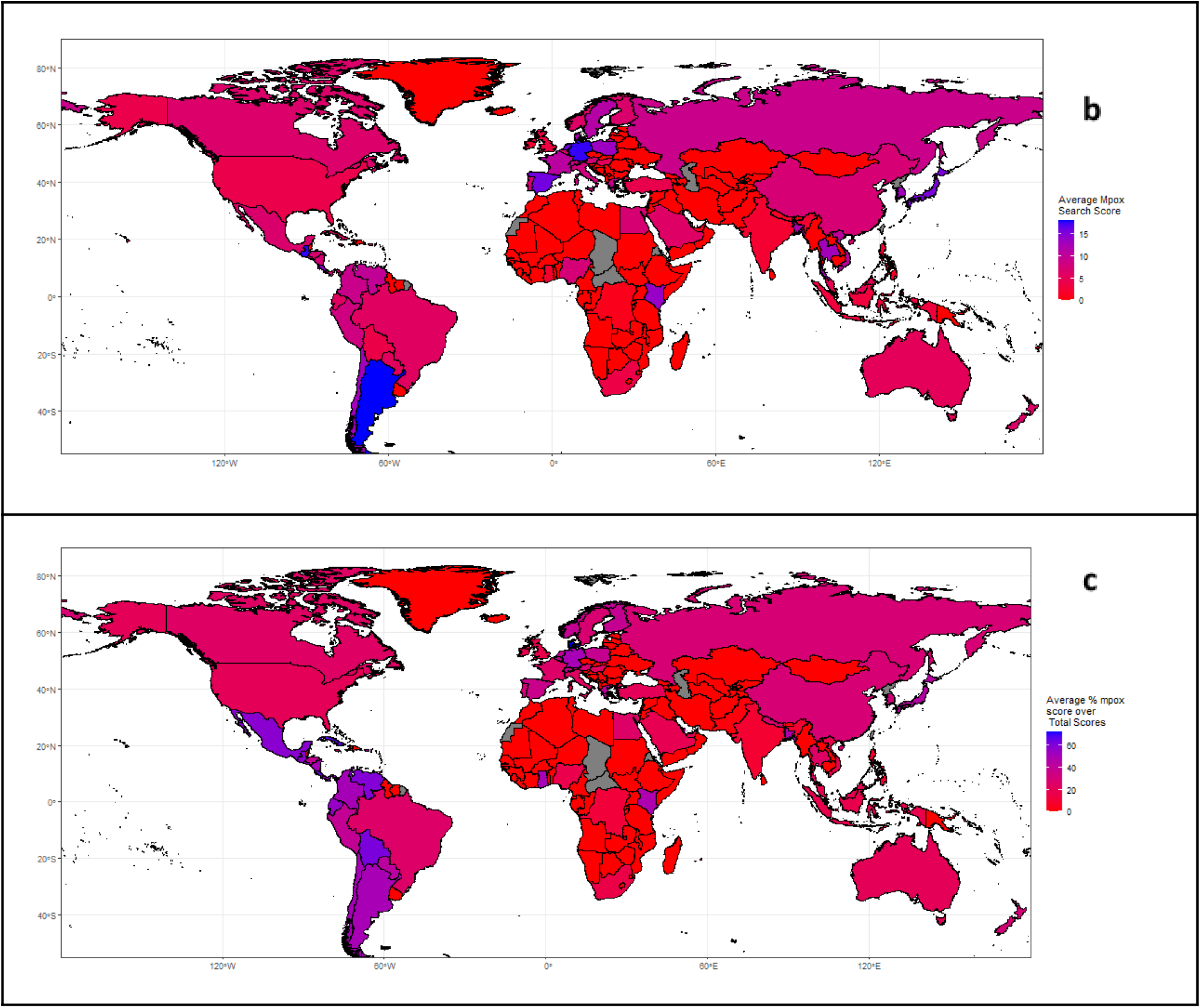
Average search score per country between 11/01/2022 and 12/01/2023. (a) Average search score for monkeypox; b) average search score for mpox; and c) difference between mpox and monkeypox where blue indicates higher use and red indicates lower use. Locations in gray were excluded from analyses either to missing search data or covariate data, or both.

Figure 1 above demonstrates some strong regional patterns in the search pattern for mpox with Europe and South America tending to have higher values of searches while most of Africa and the Middle East had lower search proportions; monkeypox, in contrast, showed more regional heterogeneity.

**Figure 2:**
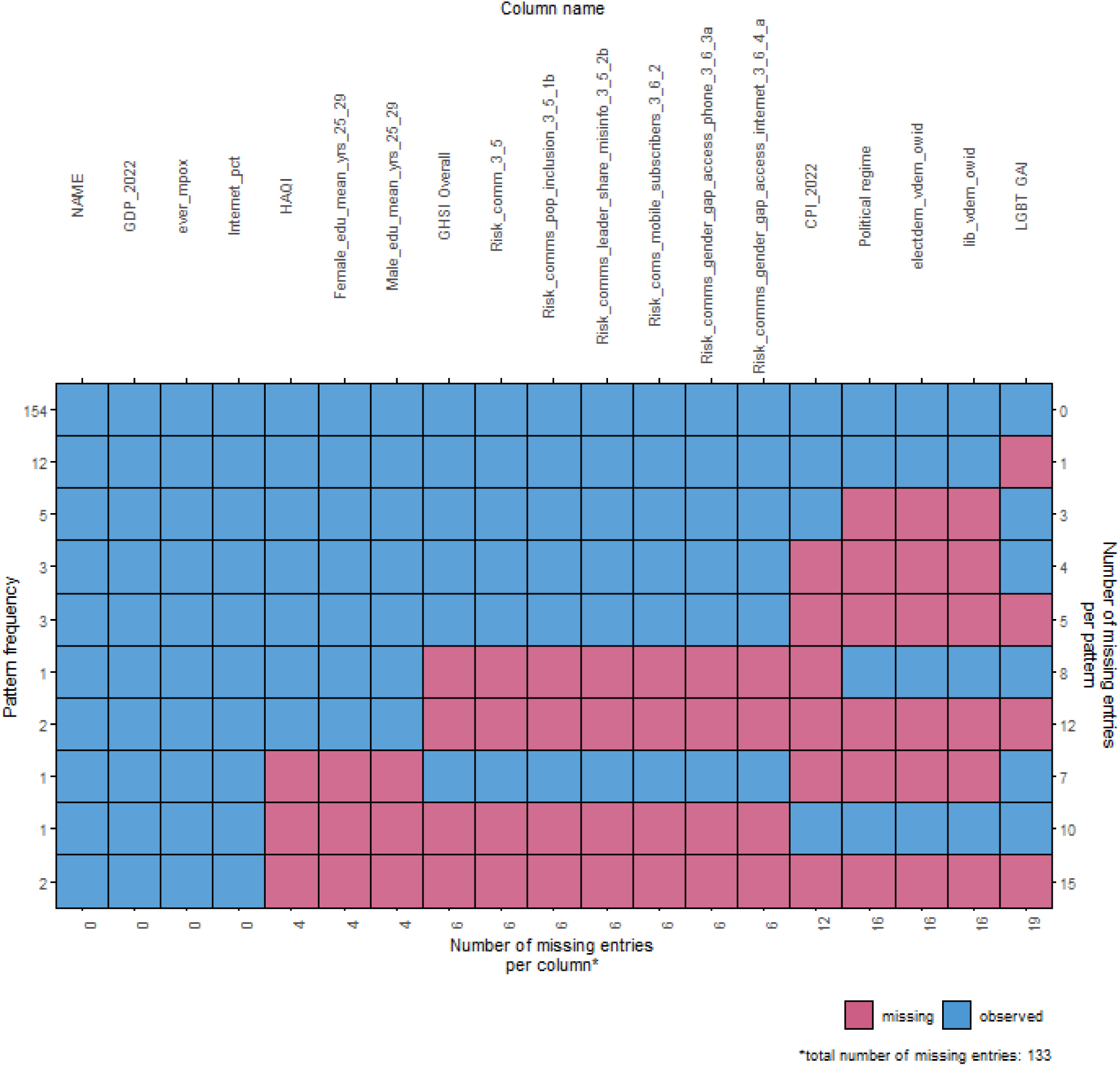
Degree of missingness for variables in the analysis.

In Figure 2, we see the missingness of our covariates across the entire dataset. 154 observations had all covariate data, another 12 were only missing GAI, and the remaining 18 entries were missing multiple covariates.

**Figure 3:**
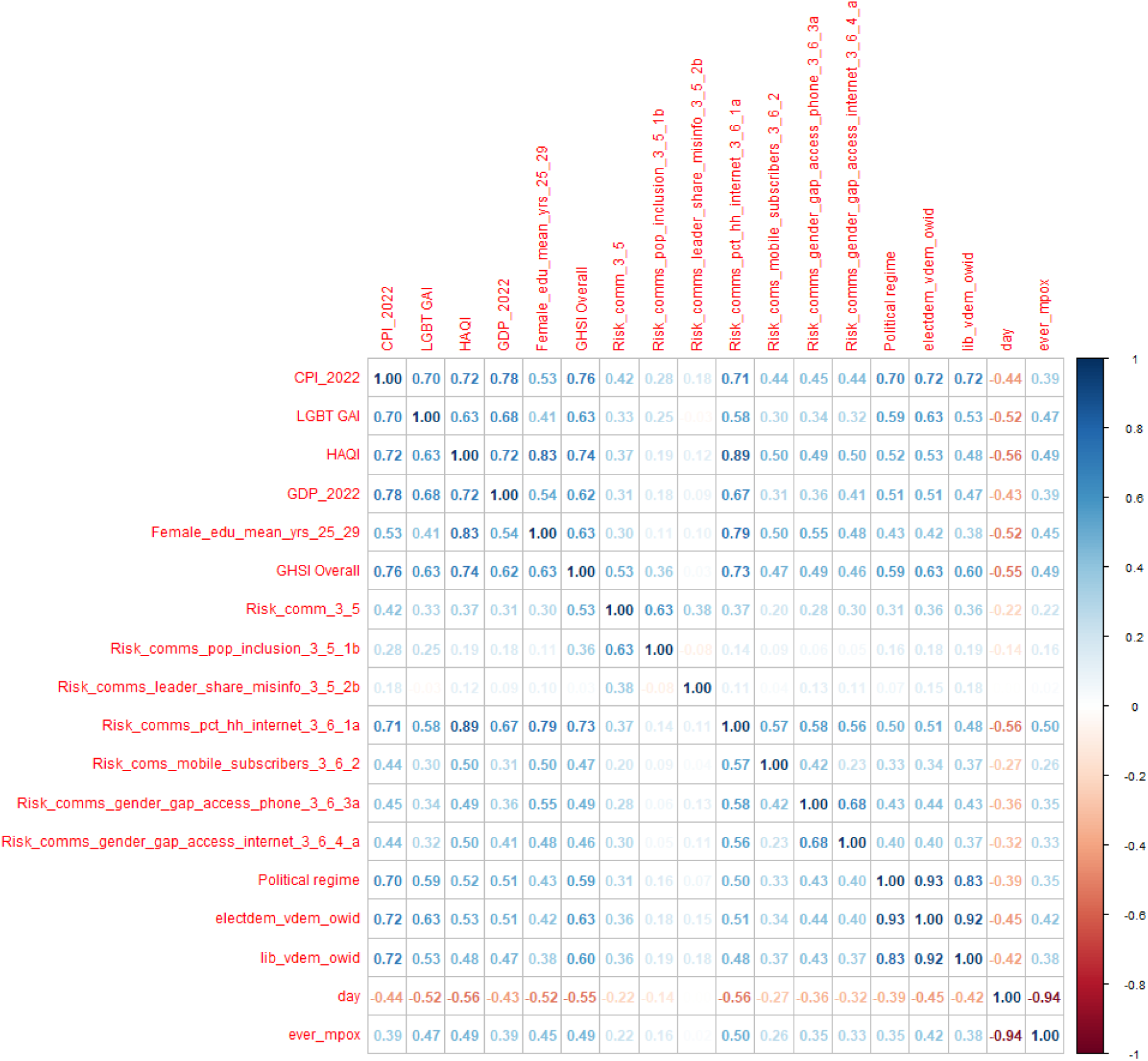
Correlation between all covariates evaluated in univariable model.

Figure 3 above shows the correlations between all the covariates included in the univariable model. Several covariates demonstrate high multicollinearity.

**Figure 4:**
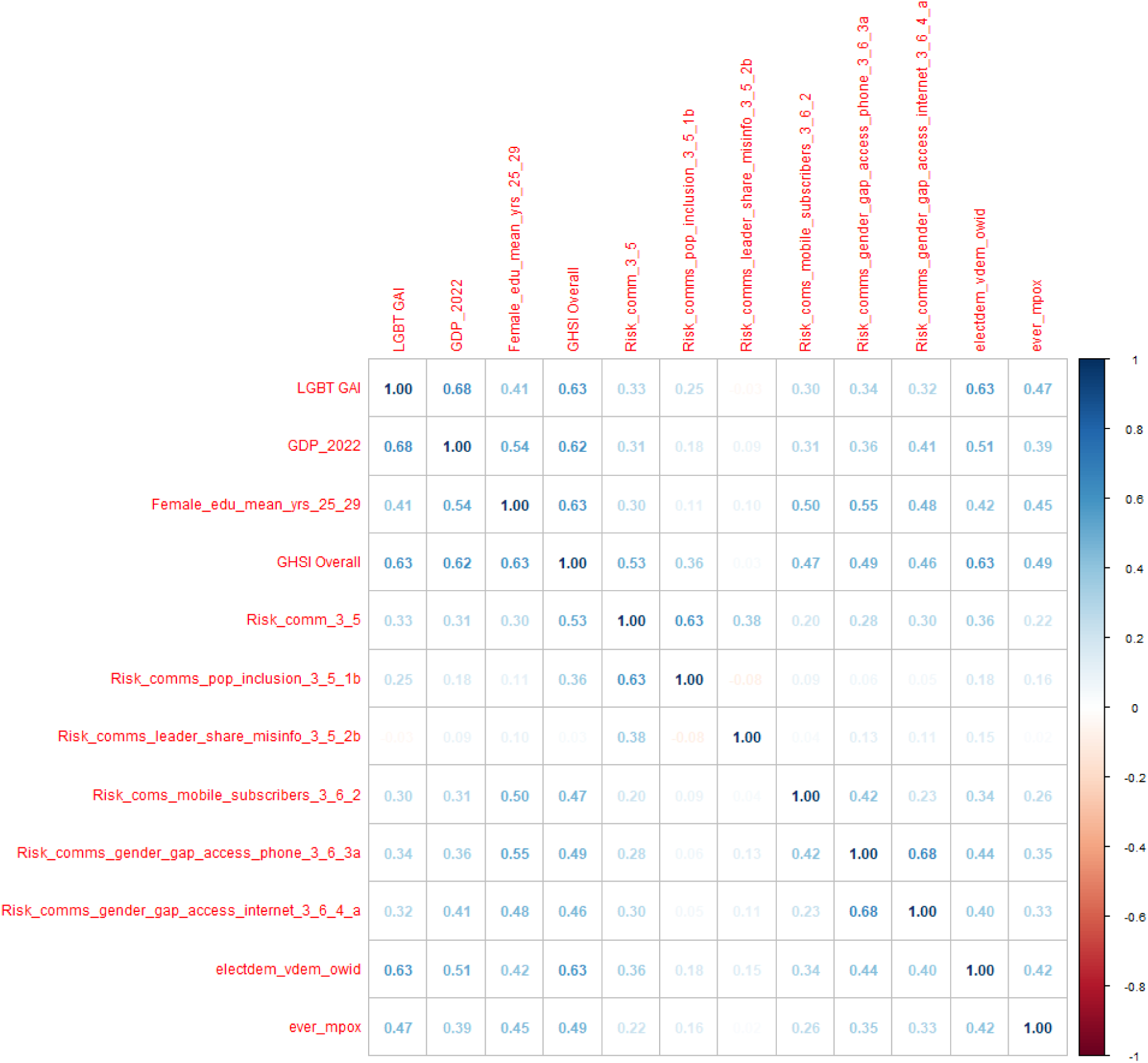
Correlation between all included multivariable model covariates.

In contrast to Figure 3, Figure 4 shows the collinearity for only those variables included in the final multivariable model. The highest correlation observed was 0.63 versus −0.94 in Figure 3.

**Figure 5:**
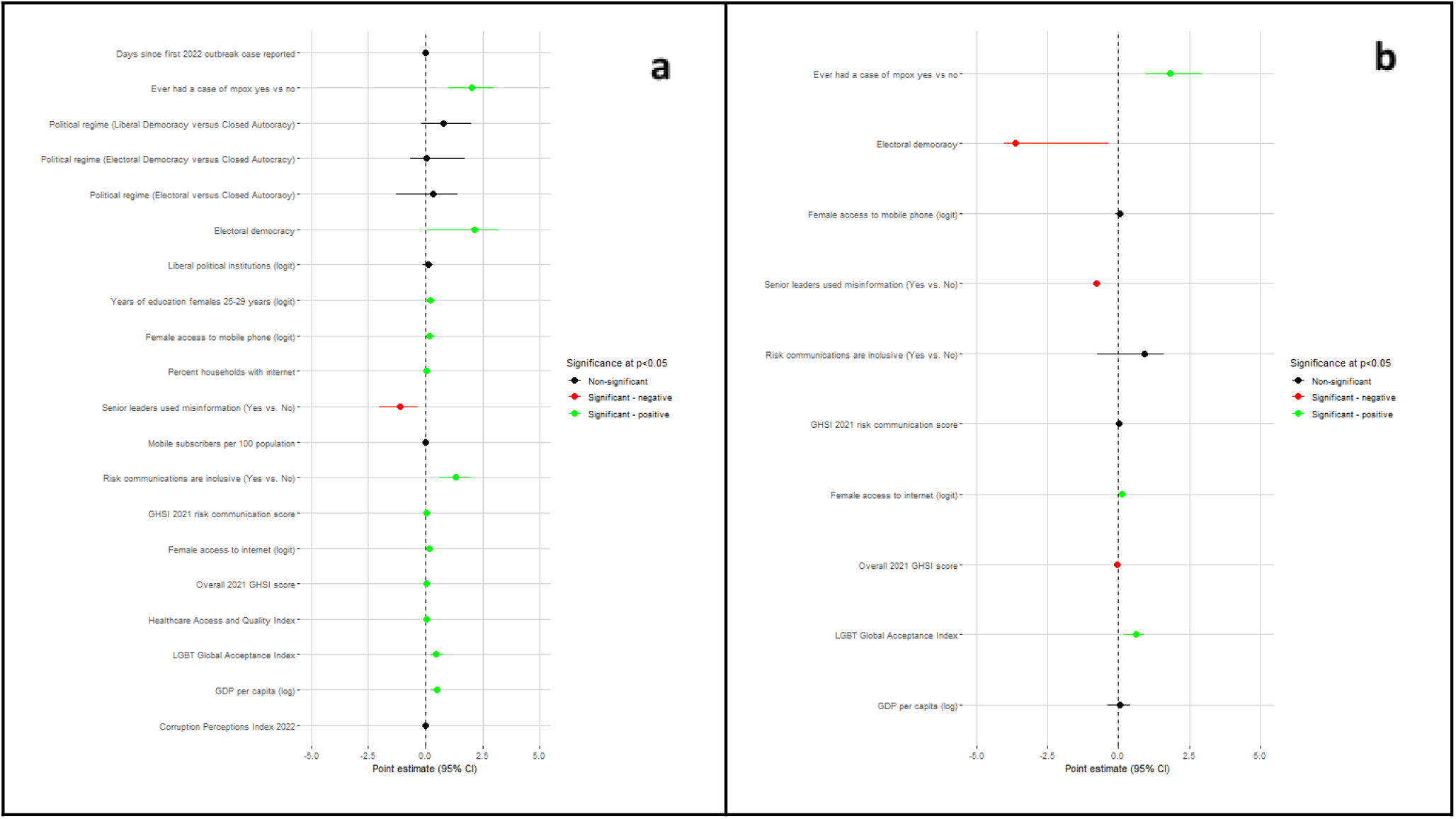
Country-level factors associated with ‘mpox’ adopter countries (binary outcome) in univariable (a) and multivariable (b) models in native language of UN languages. Variables that were statistically significantly associated with the binary outcome of interest at alpha = 0.1 in the corresponding univariable models were included in the multivariable model. Highly collinear variables were removed before fitting the multivariable model.

In Figure 5, we see 13 significant associations in the univariable model. The multivariable model suggests a significant positive association with ever having had a case of mpox, LGBTQ+ Global Acceptance Index, and ratio of female-to-male access to the Internet and a significant negative association with electoral democracy, senior leaders having propagated misinformation, and 2021 overall GHSI score.

**Figure 6:**
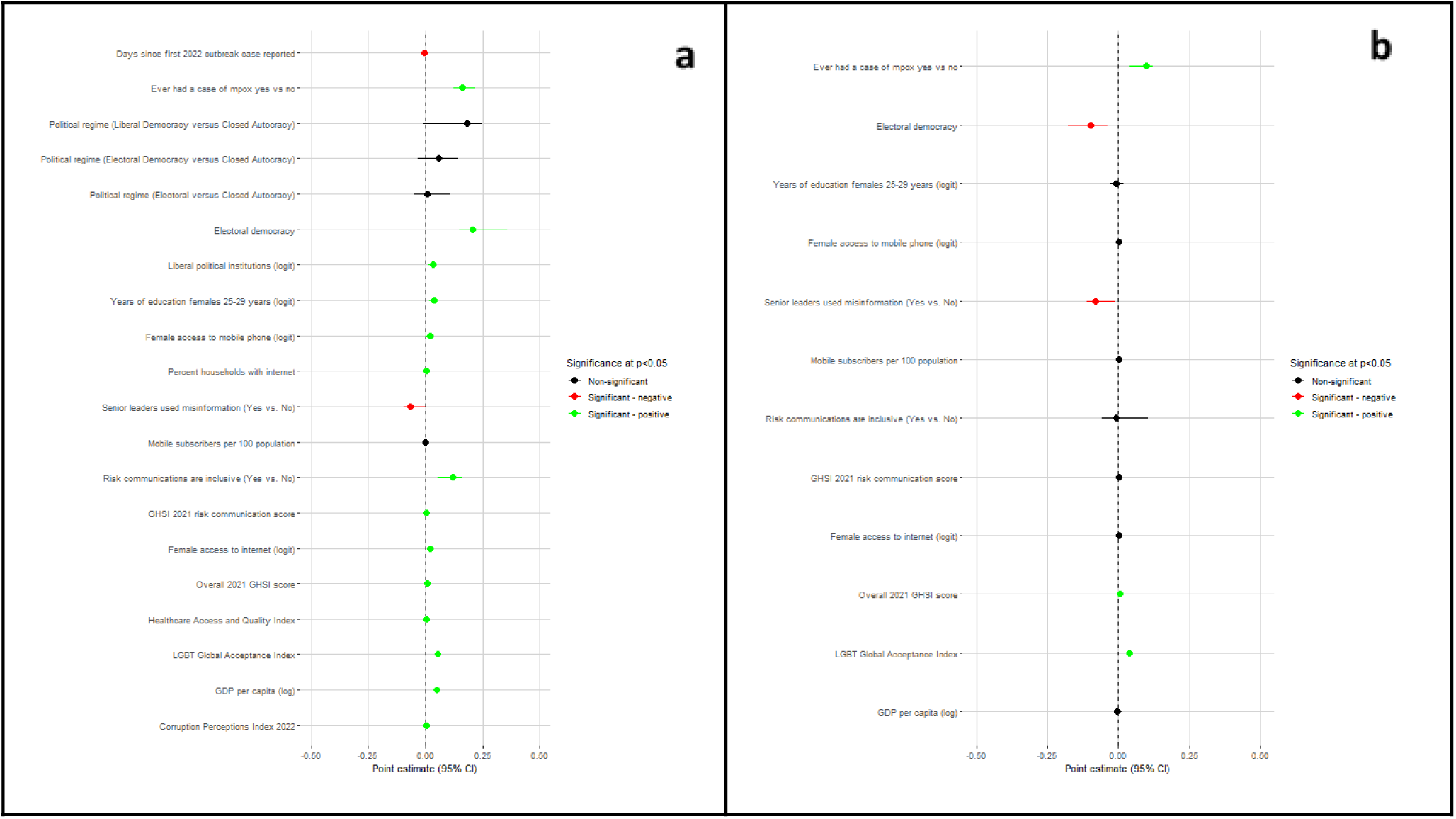
Country-level factors associated with ‘mpox’ search proportion (continuous outcome) in univariable (a) and multivariable (b) models, in native language of UN languages. Variables that were statistically significantly associated with the continuous outcome of interest at alpha = 0.1 in the corresponding univariable models were included in the multivariable model. Highly collinear variables were removed before fitting the multivariable model.

In Figure 6, we see 16 significant associations in the univariable model. The multivariable model suggests a significant positive association with ever having had a case of mpox, LGBTQ+ Global Acceptance Index, and 2021 overall GHSI score and a significant negative association with electoral democracy and senior leaders having propagated misinformation.

**Figure 7:**
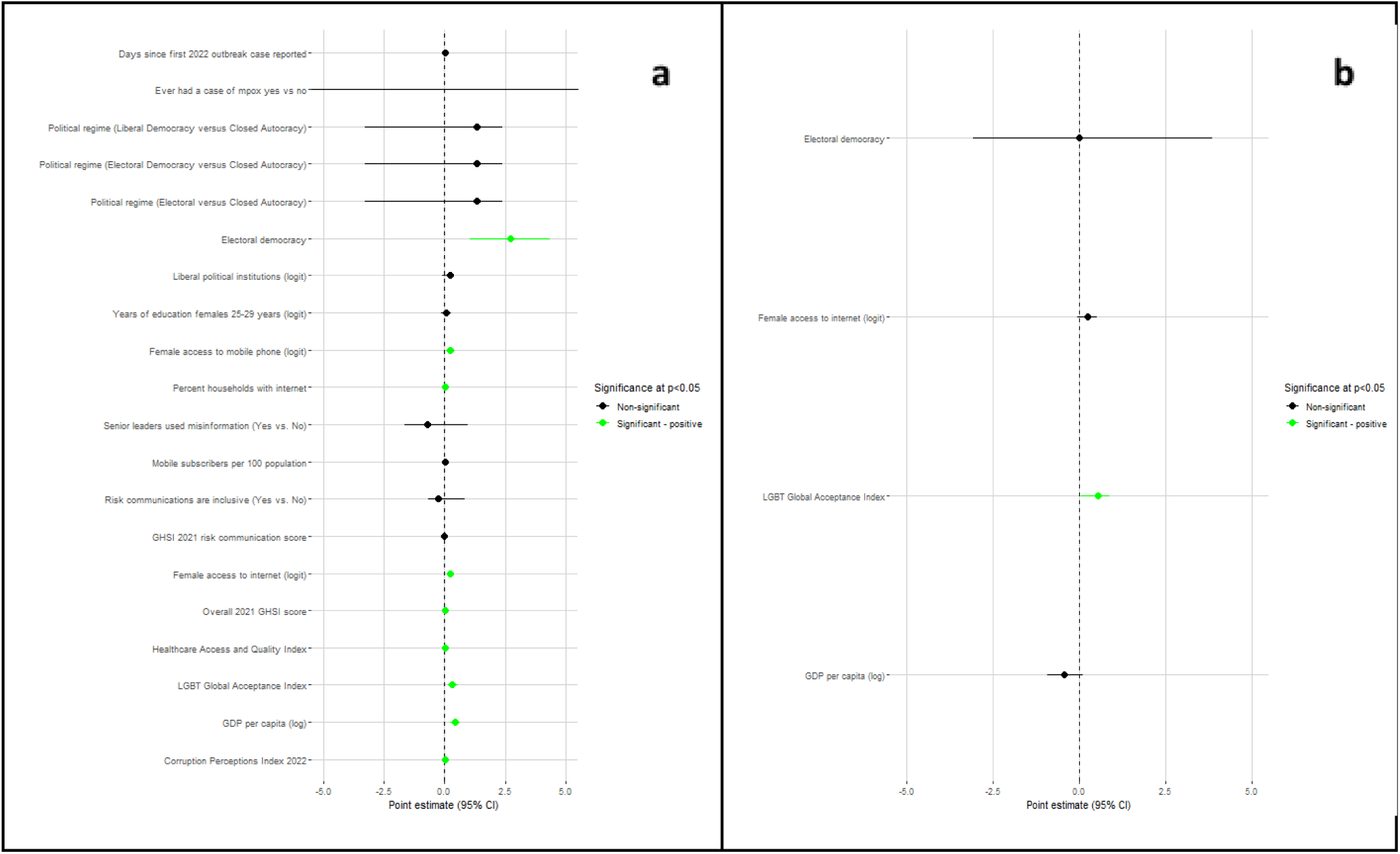
Country-level factors associated with ‘mpox’ adopter countries (binary outcome) in univariable (a) and multivariable (b) models, complete case analysis (Approach 1, n=154) Variables that were statistically significantly associated with the binary outcome of interest at alpha = 0.1 in the corresponding univariable models were included in the multivariable model. Highly collinear variables were removed before fitting the multivariable model.

In Figure 7, we see 9 significant associations in the univariable model. The multivariable model only suggests a significant positive association with LGBTQ+ Global Acceptance Index.

**Figure 8:**
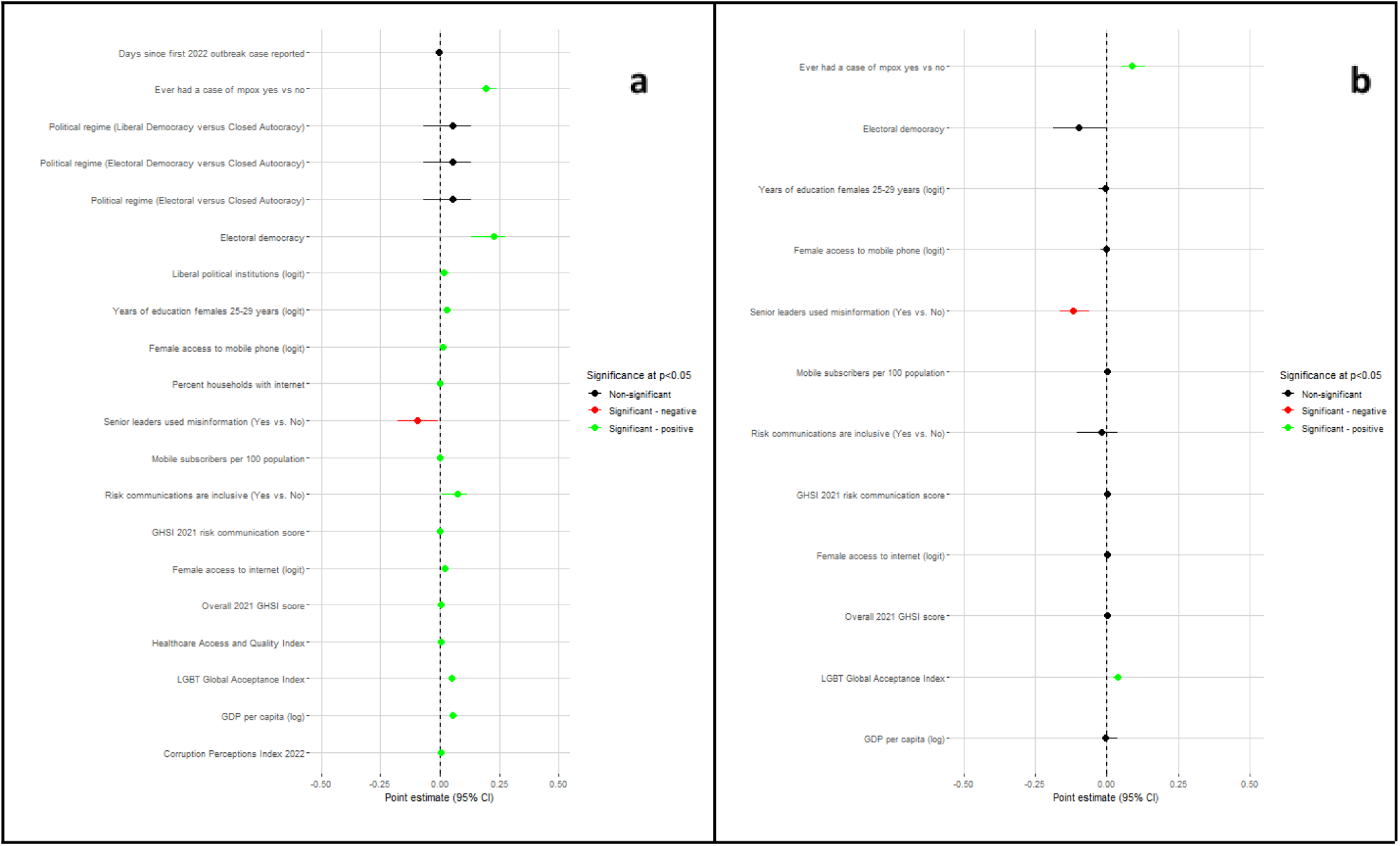
Country-level factors associated with ‘mpox’ search proportion (continuous outcome) in univariable (a) and multivariable (b) models, complete case analysis (Approach 1, n=154) Variables that were statistically significantly associated with the continuous outcome of interest at alpha = 0.1 in the corresponding univariable models were included in the multivariable model. Highly collinear variables were removed before fitting the multivariable model.

In Figure 8, we see 16 significant associations in the univariable model. The multivariable model suggests a significant positive association with ever having had a case of mpox and LGBTQ+ Global Acceptance Index and a significant negative association with senior leaders having propagated misinformation.

**Figure 9:**
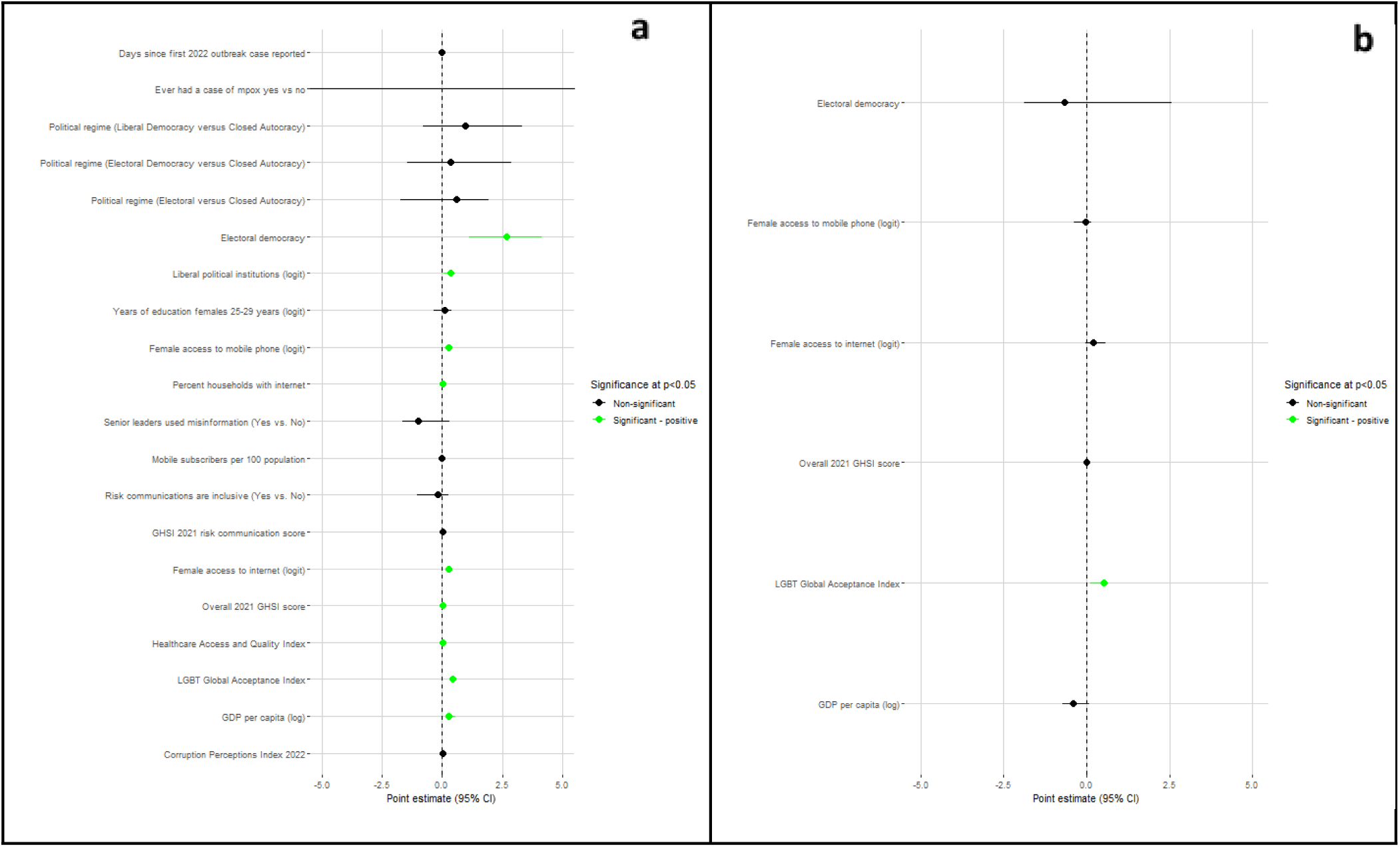
Country-level factors associated with ‘mpox’ adopter countries (binary outcome) in univariable (a) and multivariable (b) models, imputing only LGBT GAI (Approach 2, n=166) Variables that were statistically significantly associated with the binary outcome of interest at alpha = 0.1 in the corresponding univariable models were included in the multivariable model. Highly collinear variables were removed before fitting the multivariable model.

In Figure 9, we see 9 significant associations in the univariable model. The multivariable model only suggests a significant positive association with LGBTQ+ Global Acceptance Index.

**Figure 10:**
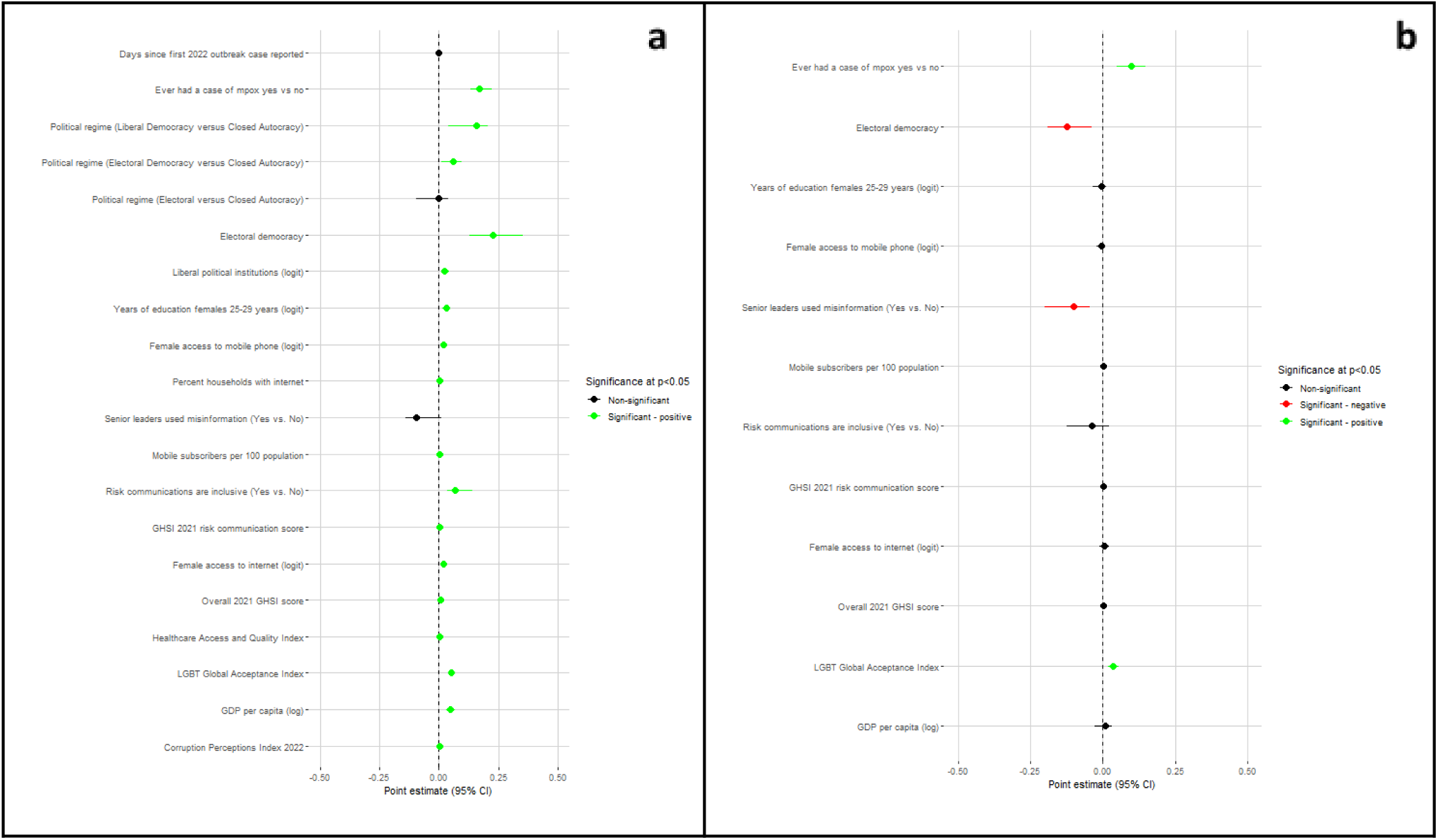
Country-level factors associated with ‘mpox’ search proportion (continuous outcome) in univariable (a) and multivariable (b) models, imputing only LGBT GAI (Approach 2, n=166) Variables that were statistically significantly associated with the continuous outcome of interest at alpha = 0.1 in the corresponding univariable models were included in the multivariable model. Highly collinear variables were removed before fitting the multivariable model.

In Figure 10, we see 17 significant associations in the univariable model. The multivariable model suggests a significant positive association with ever having had a case of mpox and LGBTQ+ Global Acceptance Index and a significant negative association with electoral democracy and senior leaders having propagated misinformation.

**Figure 11:**
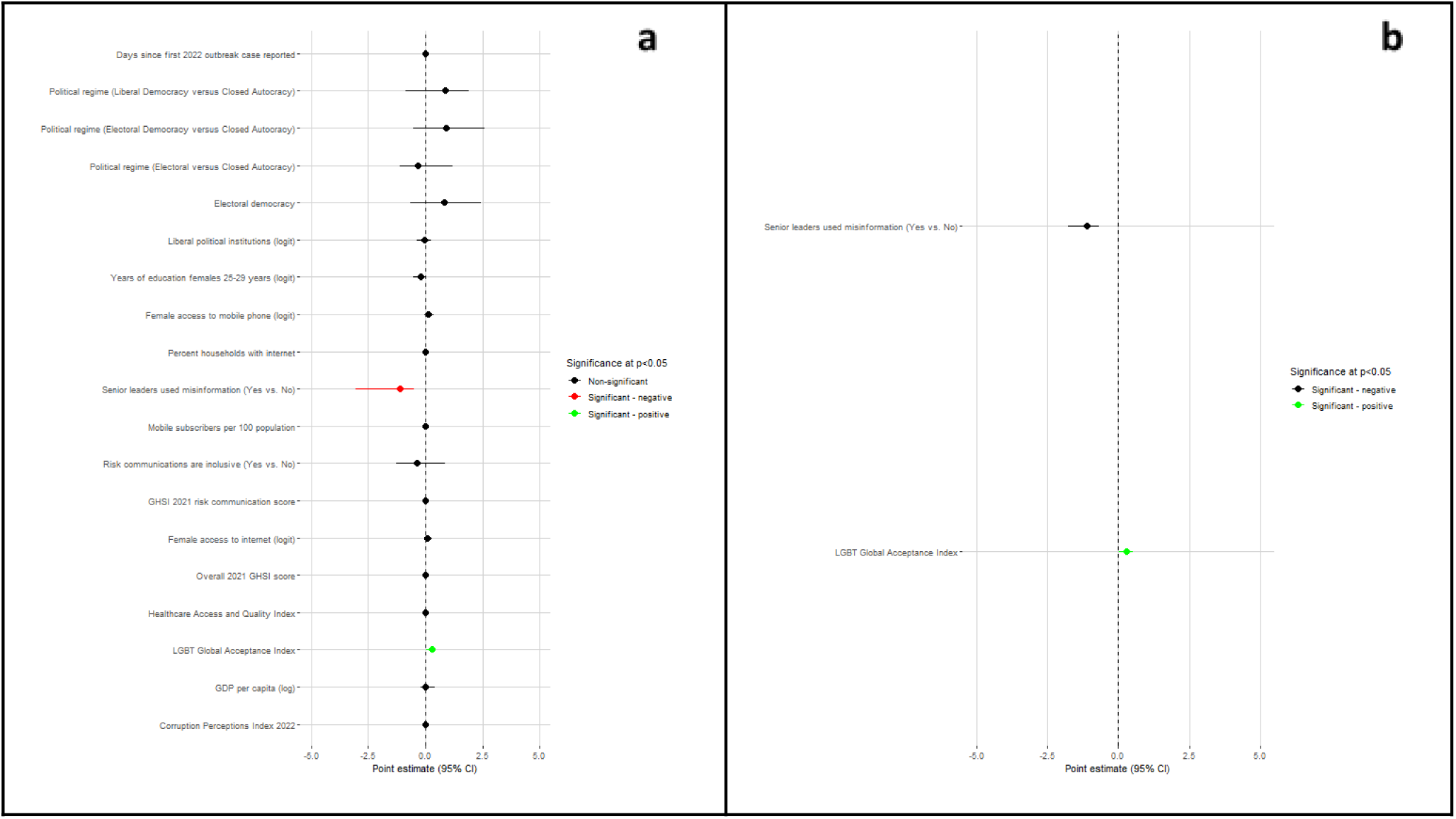
Country-level factors associated with ‘mpox’ adopter countries (binary outcome) in univariable (a) and multivariable (b) models in English, among countries that have ever had a case of mpox (since 1970s) Variables that were statistically significantly associated with the binary outcome of interest at alpha = 0.1 in the corresponding univariable models were included in the multivariable model. Highly collinear variables were removed before fitting the multivariable model.

In Figure 11, we see 2 significant associations in the univariable model.The multivariable model only suggests a significant positive association with LGBTQ+ Global Acceptance Index.

**Figure 12:**
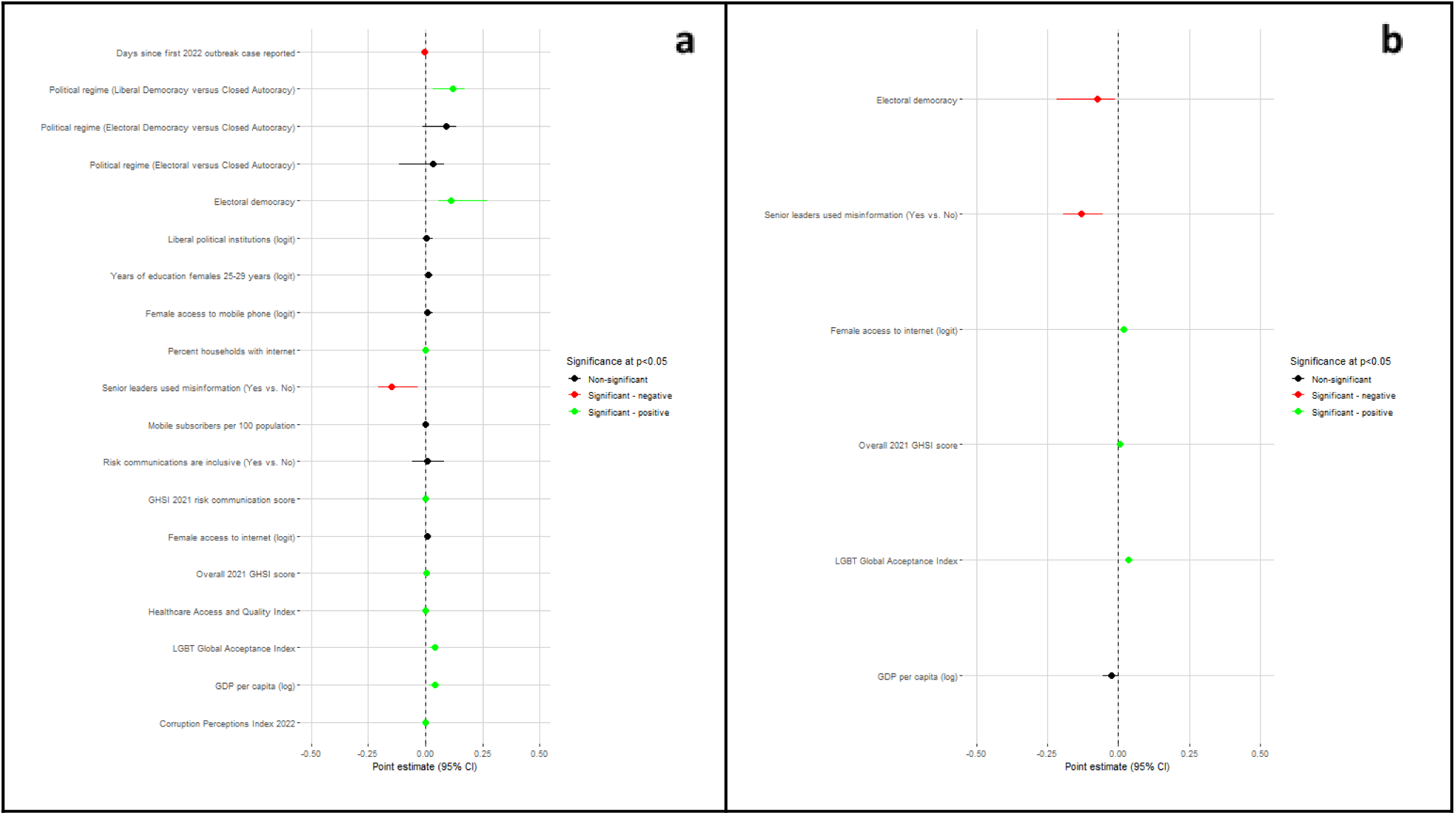
Univariable (a) and multivariable (b) model for countries with percent mpox search proportion versus total search proportion in English, among countries that have ever had a case of mpox (since 1970s) Variables that were statistically significantly associated with the continuous outcome of interest at alpha = 0.1 in the corresponding univariable models were included in the multivariable model. Highly collinear variables were removed before fitting the multivariable model.

In Figure 10, we see 11 significant associations in the univariable model. The multivariable model suggests a significant positive association with ratio of female-to-male access to the Internet, 2021 overall GHSI score, and LGBTQ+ Global Acceptance Index and a significant negative association with electoral democracy and senior leaders having propagated misinformation.

**Figure 13:**
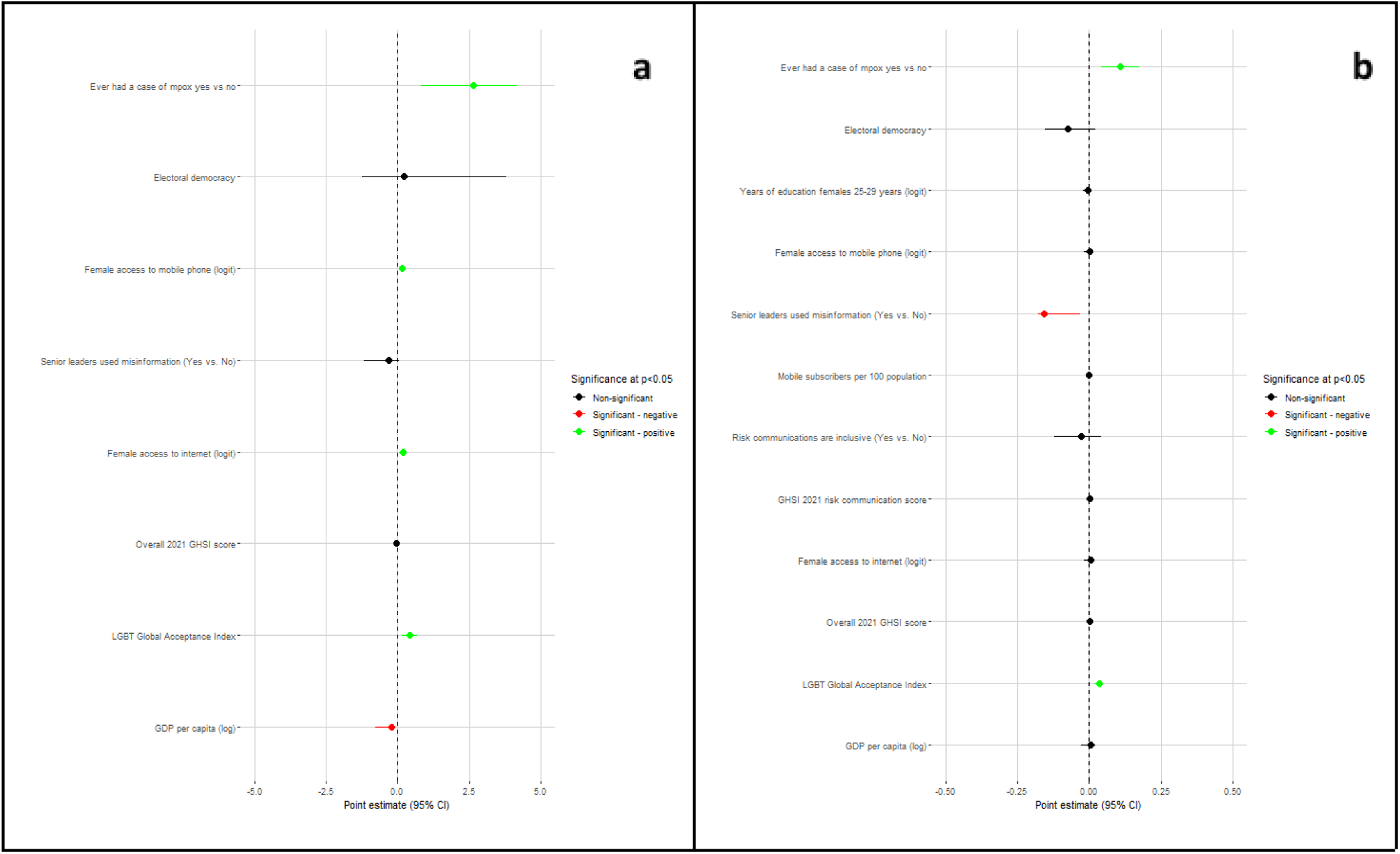
Using a threshold of 0.05 for multivariable model inclusion, binary (a) and continuous (b), full imputation (n=184) Variables that were statistically significantly associated with either outcome of interest at alpha = 0.1 in the corresponding univariable models were included in the multivariable model. Highly collinear variables were removed before fitting the multivariable model.

In Figure 13, we see the multivariable model results only for using 0.05 inclusion criteria into the multivariable model rather than 0.1. In panel a) which displays the binary model results, we see a significant positive relationship with ever having had a case of mpox, LGBTQ+ Global Acceptance Index score, ratio of female-to-male access to the Internet, and ratio of female-to-male access to mobile phones and a significant negative relationship with GDP per capita. In panel b) which displays the continuous model results, we see a significant positive relationship with ever having had a case of mpox andLGBTQ+ Global Acceptance Index score and a significant negative relationship with senior leaders having propagated misinformation.

**Figure 14:**
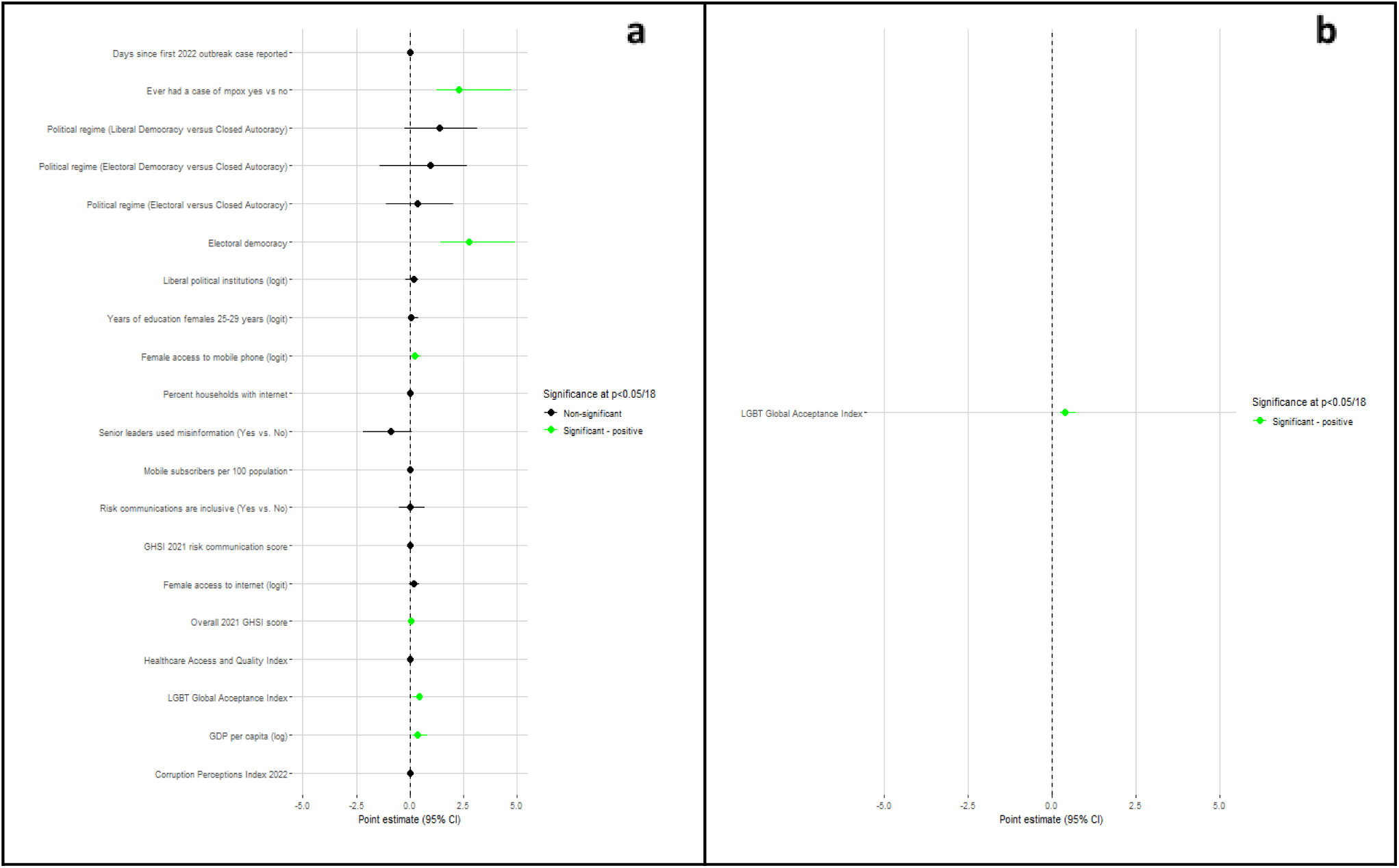
Country-level factors associated with ‘mpox’ adopter countries (binary outcome) in univariable (a) and multivariable (b) models, fully imputed using a Bonferonni correction for 18 unique variables (n=184 observations) Inclusion to multivariable is any variable significant at 0.056, removing highly collinear variables

In Figure 14, we see 6 significant associations in the univariable model.The multivariable model only suggests a significant positive association with LGBTQ+ Global Acceptance Index.

**Figure 15:**
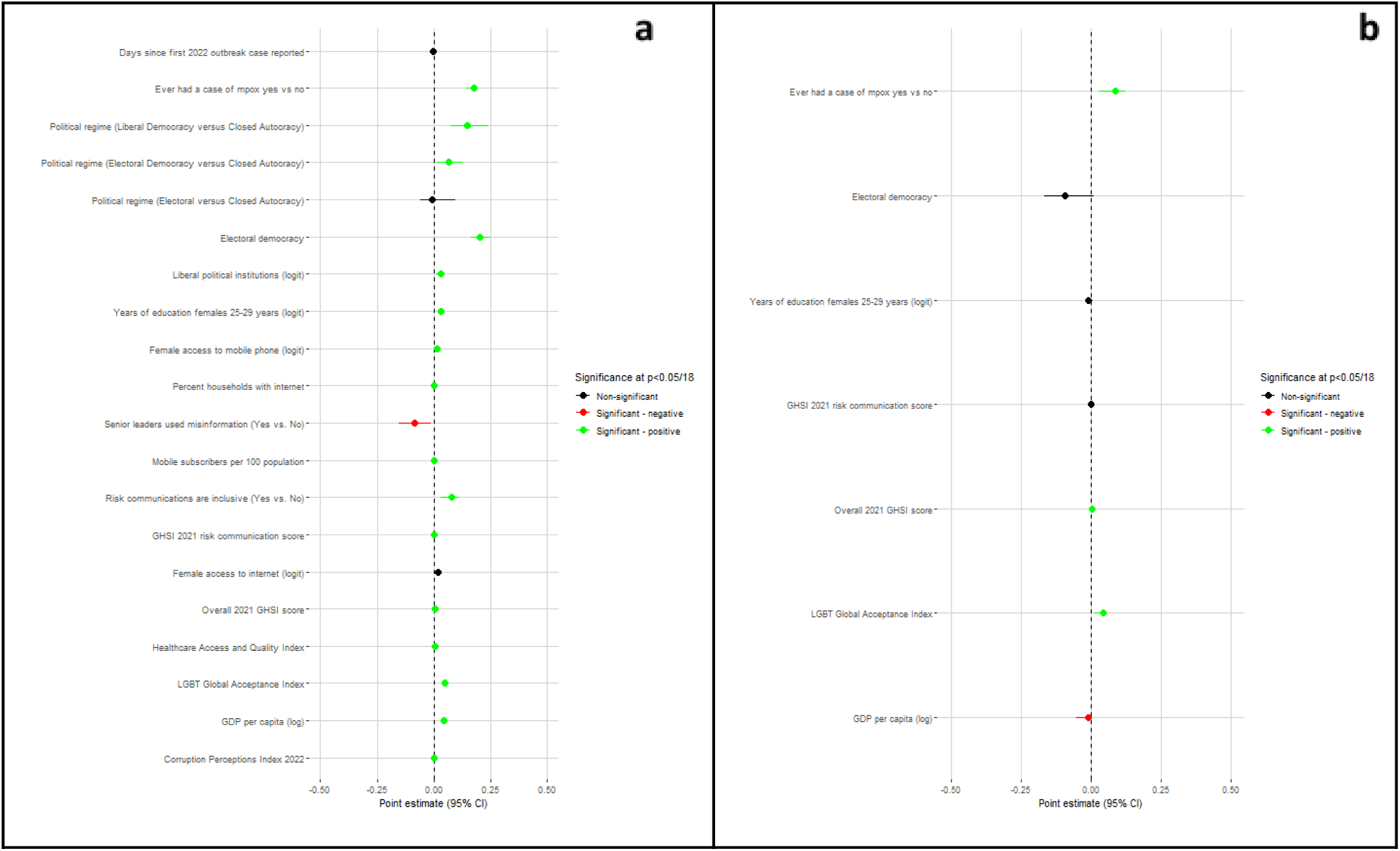
Country-level factors associated with ‘mpox’ search proportion (continuous outcome) in univariable (a) and multivariable (b) models, fully imputed using a Bonferonni correction for 18 unique variables (n=184 observations) Inclusion to multivariable is any variable significant at 0.056, removing highly collinear variables

In Figure 15, we see 17 significant associations in the univariable model. The multivariable model suggests a significant positive association with ever having had a case of mpox, LGBTQ+ Global Acceptance Index, and 2021 overall GHSI score and a significant negative association with GDP per capita.

**Figure 16:**
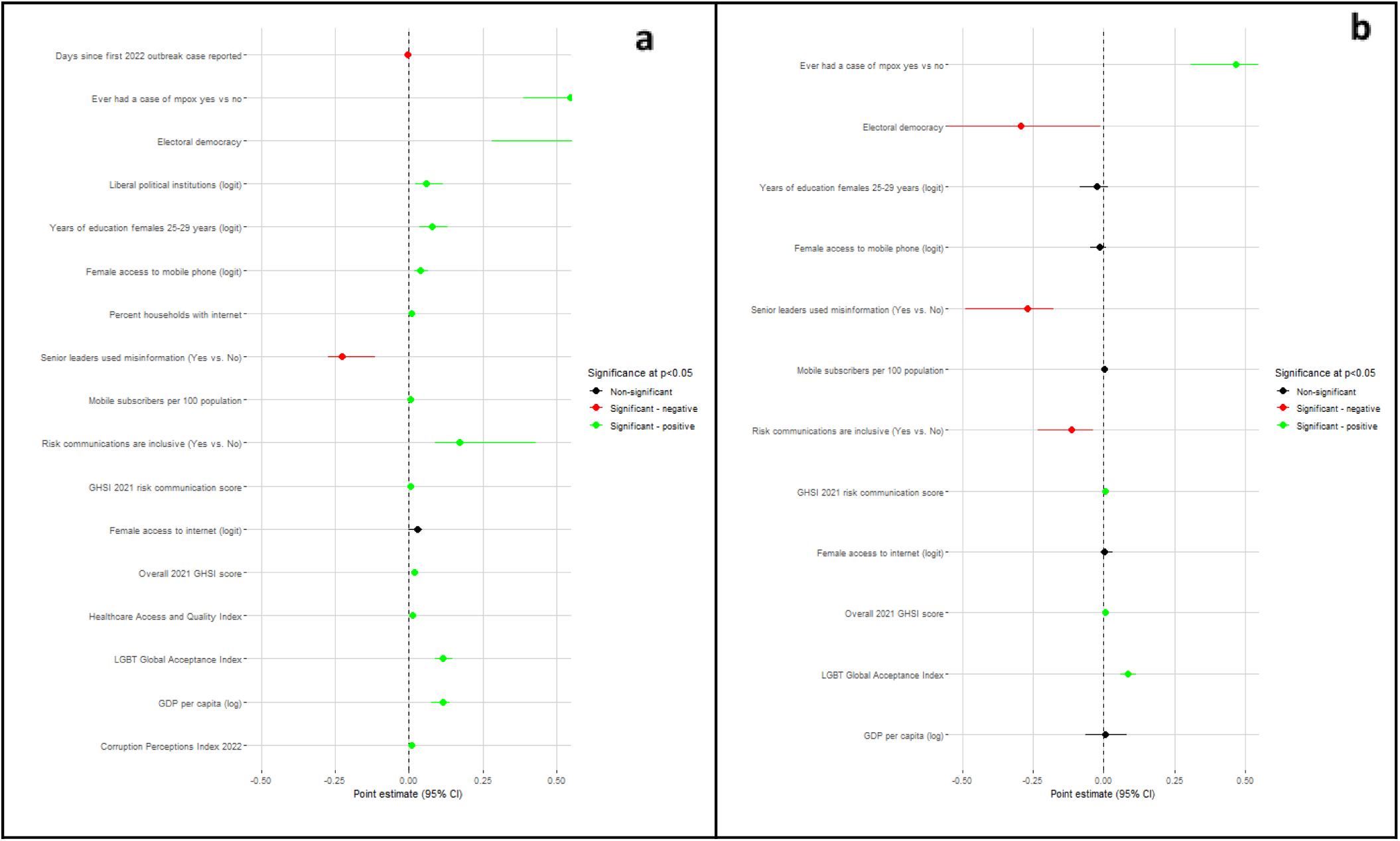
Country-level factors associated with ‘mpox’ search proportion (continuous outcome) in univariable (a) and multivariable (b) models, fully imputed using a Tobit model for 17 unique variables (n=184 observations) Variables that were statistically significantly associated with the binary outcome of interest at alpha = 0.1 in the corresponding univariable models were included in the multivariable model. Highly collinear variables were removed before fitting the multivariable model.

In Figure 16, we see 16 significant associations in the univariable model. The multivariable model suggests a significant positive association with ever having had a case of mpox, LGBTQ+ Global Acceptance Index, 2021 overall GHSI score, and 2021 GHSI Risk Communication score and a significant negative association with electoral democracy, senior leaders having propagated misinformation, and 2021 GHSI inclusive risk communications.

**Figure 17:**
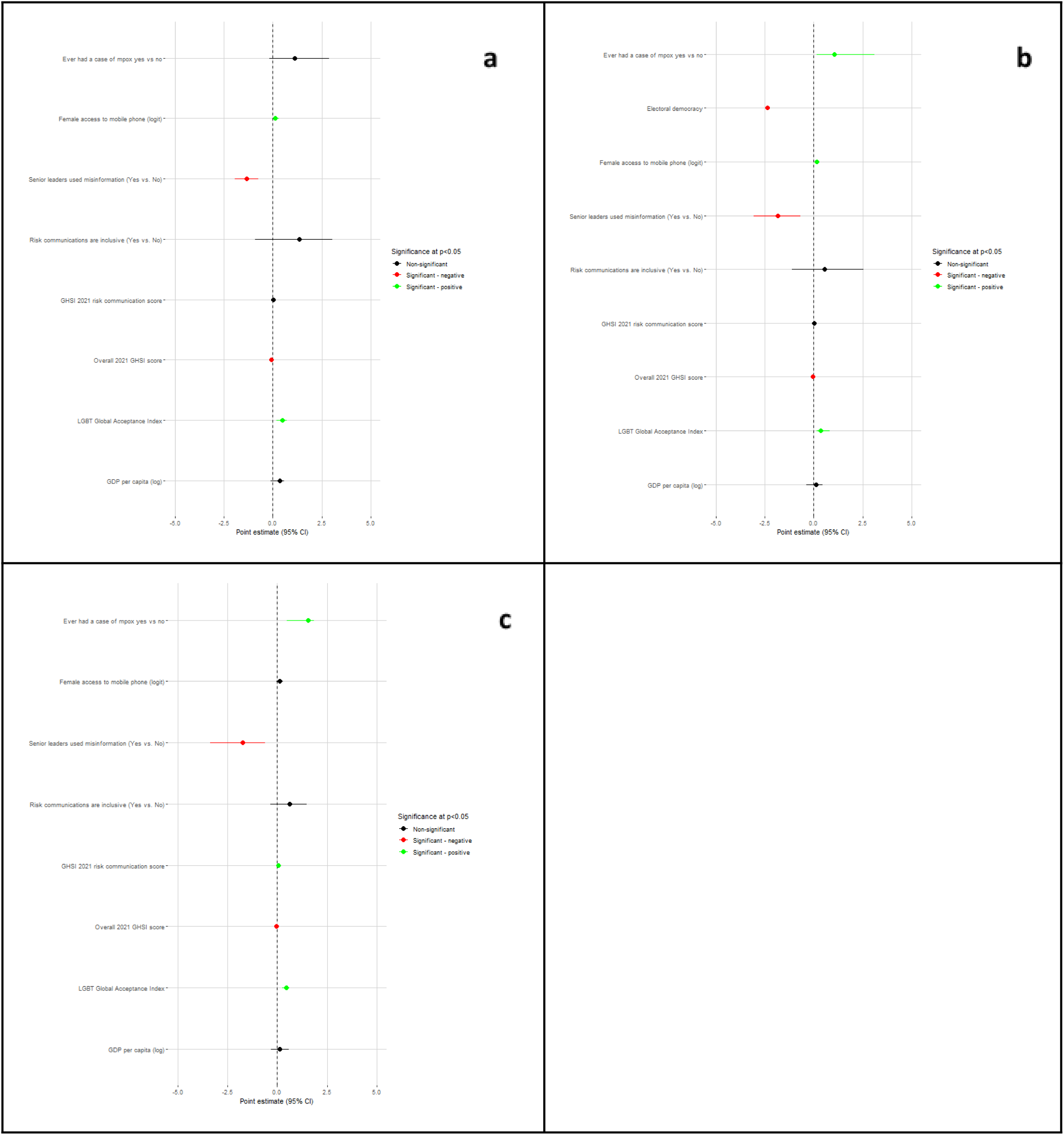
Counterfactual scenario where (a) Ecuador and Madagascar both had lower mpox search proportion than monkeypox, (b) Ecuador and Madagascar both had higher mpox search proportion than monkeypox, and (c) Ecuador and Madagascar each had opposite values than the original English-language analysis, continuous outcome, full imputation, multivariable model (n=184) Variables that were statistically significantly associated with the binary outcome of interest at alpha = 0.1 in the corresponding univariable models were included in the multivariable model. Highly collinear variables were removed before fitting the multivariable model.

In Figure 17, we see the results of our 3 counterfactual analyses. In the first (panel a), we see the results of setting both Ecuador and Madagascar to 0 for our binary variable (i.e, having both have lower ‘mpox’ searches than ‘monkeypox’ searches). This multivariable model suggests a significant positive association with LGBTQ+ Global Acceptance Index and ratio of female-to-male access to mobile phones and a significant negative relationship with senior leader propagation of misinformation and 2021 overall GHSI score.

In the second (panel b), we see the results of setting both Ecuador and Madagascar to 1 for our binary variable (i.e, having both have higher ‘mpox’ searches than ‘monkeypox’ searches). This multivariable model suggests a significant positive association with ever having had a case of mpox, LGBTQ+ Global Acceptance Index, and ratio of female-to-male access to mobile phones and a significant negative relationship with electoral democracy, senior leader propagation of misinformation, and 2021 overall GHSI score.

In the third (panel c), we see the results of setting each Ecuador and Madagascar to the opposite value they had in the main analysis (i.e, a value of 0 for Ecuador and 1 for Madagascar). This multivariable model suggests a significant positive association with ever having had a case of mpox, LGBTQ+ Global Acceptance Index, and 2021 GHSI risk communication score and a significant negative relationship with senior leader propagation of misinformation, and 2021 overall GHSI score.

## Section 5: Summary of Sensitivity Analyses

Our sensitivity analyses largely showed consistent findings with our main outcomes. For our binary outcome of being an ‘mpox’ adopter country (i.e., searching for ‘mpox’ more than ‘monkeypox’), all six sensitivity analyses (using other UN languages, focus only on countries with a history of mpox, complete case analysis, imputing GAI only, using 0.05 as the multivariable model threshold, and the Bonferroni model, Figures 5, 7, 9, 11, 13a, and 14) showed a significant, positive relationship with the LGBTQ+ GAI index. For the continuous model —the ‘mpox’ search proportion — all seven sensitivity analyses (Figures 6, 8, 10, 12, 13b, 15, and 16) showed a significant positive relationship with both the LGBTQ+ GAI index; similarly, all six sensitivity analyses that included history of mpox cases were significantly, positively associated with this outcome as well. Nearly all (n=6) of the ‘mpox’ search proportion models also showed a significant, negative relationship with leaders propagating misinformation about infectious diseases. Our binary outcome was statistically associated with ever having had a case of ‘mpox’ in n=2 models and senior leaders having propagated misinformation in n=1 models.

In turning to secondary results, the majority (n=4) of ‘mpox’ search proportion models showed a significant positive relationship with the 2021 overall GHSI score and a significant negative relationship with electoral democracy. Additional variables that were occasionally significant with our binary model, a positive relationship with the ratio of ratio of female-to-male access to each the Internet (n=2) and a mobile phone (n=1), a negative relationship with 2021 overall GHSI score (n=1), a negative relationship with electoral democracy (n=1), and a negative relationship with GDP per capita (n=1). Additional variables that were occasionally significant with our continuous model were a positive relationship with the ratio of female-to-male access to the internet (n=1), a negative relationship with the 2021 GHSI measure on inclusive risk communications (n=1), a positive relationship with the 2021 GHSI overall risk communications score (n=1), and a negative relationship with GDP per capita (n=1).

